# “If we have water, we have money”: A qualitative investigation of the role of water in women’s economic engagement in Guatemala, Honduras, Kenya, and Zimbabwe

**DOI:** 10.1101/2025.04.30.25326747

**Authors:** Emily Ogutu, Sheela S. Sinharoy, Madeleine Patrick, Thea Mink, Alicia Macler, Loice Mbogo, Olivia Bendit, Ingrid Lustig, Jazmina Nohemí Irías, Sandra Antonio, Gladys Ramos, Alma Juarez, Carlos Daniel Sic, Erick Calderón, Héctor Salvador Peña Ramírez, Jorge Lemus, Everlyne Atandi, Peter Mwangi, Paul Ruto, Rohin Otieno Onyango, Gilbert Mushangari, Jammaine Jimu, Morris Chidavaenzi, Makaita Maworera, Nobuhle Dungeni Mlotshwa, Sithandekile Maphosa, Munyaradzi Damson, Bethany A. Caruso

## Abstract

**Background:** Water is essential for life and development, and access to sufficient, safe, acceptable, physically accessible, and affordable water is a basic human right. Access to water can contribute to poverty reduction, as it can enhance agriculture and livestock production and enable engagement in other water-related economic activities. Globally, 2.2 billion people lack access to safe drinking water. Women are disproportionately affected by water scarcity, as they bear the greatest burden of water related challenges, particularly in low- and middle-income countries. Water collection can deplete women’s time and energy and jeopardize their health and wellbeing, limiting their ability to participate in economic activities. Therefore, we aimed to assess the role of water on women’s economic engagement in Guatemala, Honduras, Kenya, and Zimbabwe.

**Method:** We conducted 72 focus group discussions with women (n=38; participants=298) and men (n=34; participants=202), and 56 key informant interviews (women=33; men=23) from June to October 2023 in 25 rural communities. Qualitative tools included questions about barriers, facilitators and community perceptions of women’s economic engagement, and women’s water collection experiences. We used a modified grounded theory approach for data analysis, developed deductive and inductive codes, and used MAXQDA2020 to code and organize data.

**Results:** Women’s time was compromised by limited access to sufficient quantities of water, limiting their engagement in economic opportunities. Limited water reduced livelihood activities agriculture and livestock production that women engaged in, and constrained women’s economic resources for example payment for water or for costs associated with water access, needed for income-generating activities. Health and wellbeing, specifically, physical health issues like fatigue, exhaustion, physical injuries, and pain, from water collection work, left women depleted of energy needed for economic engagement activities. Gender norms shaped roles and responsibilities for men and women and ascribed water collection as the primary responsibility of women in all four countries. Environmental factors such as drought and seasonality diminished sufficient access to water, further reducing women’s time and energy, and negatively impacted livestock and agricultural productivity due to diminished pasture and reduced water supply.

**Conclusion:** Water is a prerequisite to economic engagement, especially in hard-to-reach low-resource settings. Access to water can enable livelihood opportunities, allow women to save or reallocate time, enable various economic resource options, and improve health and wellbeing, which can then facilitate women’s economic engagement. Insufficient access to water can demand arduous water collection tasks, which can impact health by causing energy depletion, increasing risk of injury, and causing mental strain. These negative health impacts, coupled with time and opportunity costs, can constrain women’s abilities to engage in other facets of life, including economic engagement which could provide well-being benefits to women and their families. Further research is needed to understand how health and wellbeing can influence economic engagement, as many studies focus on health outcomes rather than the reverse.

## 1.0 Introduction

Water is essential for life(1) and development, and access to sufficient, safe, acceptable, physically accessible, and affordable water is a basic human right(2). Access to water can contribute to poverty reduction, as it can enhance agriculture and livestock production and enable engagement in other water-related economic activities(3, 4). However, 2.2 billion people globally still lack access to safely managed drinking water services, defined as drinking water that is from sources located on premises, free from contamination, and available when needed(5).

Inequalities in the accessibility, availability, quantity, and quality of water disproportionately affect women. Gender norms, defined here as “societal expectations that dictate acceptable behaviors, appearances, and attitudes for individuals, influencing their access to resources and freedoms, and shaping their voice, power, and sense of self” (6, 7), dictate roles and responsibilities associated with water provision, expected use and division of water in the household(5). For example, among households that need to collect water from sources located off premises, 70% of that labor is borne by women and girls globally(5). Once at water sources, women collecting water for domestic needs may be deprioritized if men arrive to provide water for livestock. In Kenya, for example, women in rural Marsabit would cope with the prioritization of men’s water needs at sources by waiting longer, going farther, paying for other sources, or collecting less water than needed(8). Furthermore, when water quantities are limited, women may ration their water use and compromise their own needs, including for drinking and hygiene, so that the needs of other household members and chores like cooking are met(9).

Water scarcity can exacerbate the time and energy burden for water collection, substantially affecting time allocation for engagement in other activities(10). Time burden refers to the time spent moving to the water source, collecting water, and returning from the water source(11) and can also include other water-related work, such as water treatment and storage. Globally, women spend an estimated 250 million hours per day collecting water and allocate significantly more time to tasks associated with water, for example laundry, than men do (12). Recent research quantifying both time and caloric expenditure of water collection in Guatemala, Honduras, Kenya, and Zimbabwe found that some women spent as many as 4.5 hours collecting water, expending as many as 952 calories in a single journey(10). Women residing in water-scarce areas spend considerable time collecting water due to the need to travel longer distances to water points, queueing, and carrying out additional waterwork like digging in the sand to extract water at riverbed sources (11, 13–15). A study in rural Benin that analyzed the impact of public water point provision on water collection times and usage in rural areas found that even with reduced travel time to the water source, women still spent substantial time queueing at the water source(11, 15). These time and energy burdens for water collection are expected to lengthen in the future due to climate change(16), which will impact women in particular(17).

Evidence suggests that constraints on water influence physical and psychosocial health and wellbeing through exposure to illnesses, injuries, and mental strain(18, 19). These health effects can negatively affect people’s capabilities, thus minimizing their contribution to production and growth(20), and reducing their full involvement in economic activities. A scoping review on the association of water carriage and water carriers’ health found that aspects of water collection, including long distances traveled, human wildlife conflicts, water retrieval work (e.g., pumping), and heavy loads led to risks and fears of physical injury, pain, exhaustion, perinatal health problems, rape, and physical abuse to women, children and people with physical disabilities(19). A lack of reliable and affordable sources of water, long distances to water sources, queuing, water management at the household level, and fear of sexual assault during water collection may all negatively impact mental health and wellbeing, especially for women(9, 19). Lack of access to sufficient and reliable water can elevate both the physical and psychological burdens of water collection and management on women(11–13). These negative effects may lead to health complications such as chest pains, fatigue, and headaches, which can affect individuals’ ability to work and generate income(11). Compromised water quality due to source location and other activities at the source can also expose women to various health risks including urinary tract infections and helminths(21). Furthermore, research has found that poor physical and mental health in early and middle adulthood have been associated with unemployment and lower socioeconomic status throughout the lifespan(22). While the impact of water insecurity on health and wellbeing is well-documented, limited information exists that directly links improved water access to health, wellbeing and economic engagement.

Beyond the time and energy burden and potential physical and mental health impacts, women’s opportunities for economic participation may be limited by their access to sufficient and reliable quantities of water(5). In one instance, a study which examined the lived experiences of women engaging in small-scale water-intensive businesses like traditional beer brewing and coffee shops in Ethiopia reported that unreliable water supply impacted their businesses(23). However, limited data exists on the relationships, and there is a need for further research to understand how women’s water access influences their economic engagement.

Similarly, limited evidence exists that demonstrates that sustainable water access and participation in water-related projects has the potential to create space for women’s participation in economic activities (24). In their study on the role of productive water use in women’s livelihood in rural Senegal, Van Houweling et al. (2015) found that access to water for productive purposes was a critical asset for expanding and diversifying women’s livelihoods(25). However, despite efforts to improve water access and water resource infrastructure, research is lacking that demonstrates how these efforts may relieve women of unpaid water related work, save their time, or result in women’s direct engagement in economic activities.

To address these existing evidence gaps, we aimed to understand the role of water in women’s economic engagement in four diverse low-income country settings: Guatemala, Honduras, Kenya, and Zimbabwe.

## 2.0 Methods

### 2.1 Study design and research partners

This qualitative study engaged participants from communities that received the Strong Women, Strong World: Beyond Access (SWSW) program implemented by World Vision in Guatemala, Honduras, Kenya, and Zimbabwe. SWSW strives to achieve sustainable empowerment of women through integration of water, sanitation, and hygiene (WASH) and livelihoods interventions while mainstreaming issues of gender equality and social inclusion. Funded by World Vision US, Emory University was the lead learning partner for research related to SWSW in four countries where the program was delivered. To enable collaborative learning, the Emory team held workshops with members of each World Vision country team leading SWSW activities to identify priority research questions of interest. As the SWSW program theorizes that water access is fundamental to women’s abilities to engage in economic activities, country teams agreed it would be useful to assess this theory. To that end, Emory designed this qualitative research study to investigate if and how water plays a role in women’s economic engagement.

In each country, World Vision selected a local learning partner to lead data collection activities, including obtaining local ethical clearance certificates, recruitment, consenting, facilitation of interviews and discussions, translation, and data management under the guidance of the Emory University team. The local learning partners were: Centro Universitario de Occidente, Universidad de San Carlos de Guatemala, Universidad Nacional Autónoma de Honduras, St. Paul’s University (Kenya), and Datalyst Africa (Zimbabwe). Members of the Emory team trained local research partners in each country. Training (described in section 2.3) was interactive and provided opportunities for the local learning partner team members to provide feedback on the research protocol and tools.

Members of the Emory team led analysis of data collected across all four countries. To gain reflections and insights about the research findings and to facilitate team members’ broad involvement and contribution as authors on the final manuscript(26) (which has been particularly challenging among WASH researchers in low and middle income counties(27), the Emory team held interactive workshops to promote discussion around findings with members of local learning partner and WV country office teams. Discussions informed the present paper, particularly the presentation of the results and discussion.

### 2.2 Study eligibility and sampling

We carried out multi-stage sampling to select study participants. First, World Vision US purposively selected the four countries where research was to take place; these countries were eligible because they were in the process of initiating SWSW programming and had the potential to integrate research learnings into these programs going forward (SWSW programming was not being initiated in any other countries at this time). Second, the WV teams leading the SWSW programming in each country purposively selected areas where the research was to take place. Geographical areas were eligible if they had already initiated SWSW programming or were set to in the months to come. Kenya and Zimbabwe teams selected two rural areas, Guatemala teams selected three rural areas for cultural variability, and Honduras selected one rural and one peri-urban area was selected. Third, the WV teams leading SWSW programming purposively selected the specific communities for research activities within each geographical area. Communities were eligible if they were to receive SWSW programming. Consideration was given to contextual factors like diversity of water source types, geographical placement, and socio-cultural variability. The safety of those carrying out and supporting data collection was also a key consideration in community selection. In all, 25 communities across four countries were engaged (Guatemala: N=7; Honduras: N=6; Kenya: N=6; Zimbabwe; N=6); 2 (both in Kenya) had received improved water infrastructure

Within identified study communities, learning partners selected a convenience sample of individuals to participate in the qualitative research activities. Individuals were eligible to participate if they were aged 18 years and above, resided in the selected communities, and were able to provide informed consent. Prior to participant selection, local World Vision and learning partner team members engaged with community leaders to discuss the study and seek their approval for the local learning partners to engage with community members. Additionally, to recruit potential participants, community leaders worked with the local partners to identify women and men who met the eligibility criteria. The community leaders were engaged because they were familiar with their communities and understood the characteristics of the community members. Local World Vision teams were not involved in the recruitment of individual participants to minimize bias in participants’ responses. Additional information on ethical, cultural, and scientific considerations specific to inclusivity in global research is provided in S1 Checklist.

### 2.3 Data collection

Qualitative data was collected from June 2023 to October 2023 (Guatemala: July 10-August 9; Honduras: July 5-August 5; Kenya: June 21-July 24; Zimbabwe: July 14-July 22 and September 20-October 1). Focus group discussions (FGDs) and key informant interviews (KIIs) were conducted with women and men from all four countries.

FGDs. FGDs were conducted because they are well suited to capture a diversity of experiences as well as community consensus around a specific topic, while KIIs were conducted to gather in-depth information from community leaders’ perspectives across the four countries(28, 29). Specifically, FGDs with women sought to understand community norms on water, gender roles, and responsibilities; perceptions and opinions of women’s economic engagement; and women’s experiences and perspectives on their ability to engage in economic activities, including activities women are engaged in and their perceived barriers and facilitators. FGD guide for women had general probes on barriers and facilitators to economic engagement, specific probes on water collection, how water might influence economic activities, and their perceptions about how improved water access may impact their lives, including engagement with economic activities.

FGDs with men were conducted to understand community norms on water, men’s perceptions on women’s involvement in economic activities and their perspectives about women’s ability to engage in these activities. FGD guide for men asked about what activities men and women are engaged in; perceived barriers and facilitators to economic engagement, with probes on water collection; men’s perception on how improved water access may impact women’s lives, with probes on women’s engagement in economic activities; and men’s opinions about women’s engagement in economic empowerment programming.

In each country we planned to conduct at least one FGD with women and one FGD with men per community, resulting in at least 12 per country (14 in Guatemala) sample sizes considered adequate to achieve saturation(30, 31). We allowed for the possibility of adding additional FGDs if it was deemed best to break the FGDs up by age groups, depending on context. In Kenya and Zimbabwe, we held some FGDs with younger and older women and older and younger men separately; partners advised that participants would feel more comfortable in groups with similarly aged peers. In all, we conducted 72 FGDs (38 with women; 34 with men; See Table 1 for a breakdown by country). FGDs had 6-12 participants, took 60-120 minutes, and were conducted in local languages in each country; K’iche’ and Mam in Guatemala, Spanish in Honduras; Samburu in Kenya; and Shona and Ndebele in Zimbabwe.

**Table 1.** Data collection activities and participants, by country.

|  | Guatemala | Honduras | Kenya | Zimbabwe | Total Activities | Total Participants |
| --- | --- | --- | --- | --- | --- | --- |
| **FGDs with Women** | 7 | 7 | 11 | 13 | 38 |  |
| Women Participants | 54 | 64 | 80 | 100 |  | 298 |
| **FGDs with Men** | 7 | 6 | 12 | 9 | 34 |  |
| Men Participants | 34 | 39 | 80 | 49 |  | 202 |
| **KIIs with Women** | 13 | 7 | 7 | 6 | 33 | 33 |
| **KIIs with Men** | 5 | 6 | 5 | 7 | 23 | 23 |
| **Total** |  |  |  |  | 128 | 556 |
|  |  |  |  |  |  | (331 W; 225 M) |

KIIs. KIIs were conducted with local leaders, both men and women, to better understand community leaders’ experiences and perspectives related to water access and use, and their perceptions of women’s economic engagement. Interview guides had specific questions on demographic information, water access and use, economic empowerment activities and decision making related to water and economic empowerment. We aimed to conduct 50 KIIs with different informants in each country to gather in depth insights and perspectives on issues around water and economic engagement(30). We conducted 56 KIIs (36 with women; 23 with men). (See Table 1 for a breakdown by count y). KIIs were conducted in the participant’s preferred language and lasted 60-90 minutes.

Research Assistants. At least eight research assistants were engaged in data collection in each country (14 in Guatemala, 10 in Honduras, 10 in Kenya, and 8 in Zimbabwe). The research assistants 1) were from the country where data collection was conducted, 2) had an understanding of the communities where data collection was to take place, and 3) were fluent speakers of the local language. Some had some experience in qualitative data collection, but all were to be trained so previous experience was not a requirement. Prior to data collection, Emory University team members engaged the research assistants who would be collecting the data in a week-long training. Training topics covered included the overall purpose of the study, current background on water and economic engagement, qualitative research methods, ethical conduct of research, transcription and translation, debriefing, and data management. Qualitative tools were translated into local languages and validated by the research assistants to ensure accuracy of the translation and cultural appropriateness. Before data collection, the tools were piloted and adapted based on input from those who collected the data.

### 2.4 Data Management

FGDs and KIIs were audio recorded, and the audio files were uploaded on a secure file storage service certified for storage of sensitive data. Audio recordings in K’iche’, Mam, and Spanish were first transcribed into Spanish by trained members of the local research teams and then translated into English by a hired translator. Audio recordings in Samburu, Ndebele, and Shona were simultaneously transcribed and translated into English by trained team members. For quality, transcripts were reviewed against corresponding audios by a different member of the data collection team to ensure accuracy of data. All transcribed data and physical copies were uploaded in the secured service storage.

### 2.5 Data analysis

A modified grounded theory approach was used to analyze the data(32). Analysis began in the field with debriefing(33). Specifically, the field team debriefed at least once a week after data collection in a specific community, discussing themes and identifying areas which required more probing(33). After fieldwork was completed, analysis continued with memoing of data, which was organized in a Microsoft Excel sheet to facilitate weekly meetings with the Emory research team to discuss themes, including commonalities and differences in barriers and facilitators of women’s economic engagement within the communities and across the countries. A codebook was developed inductively using memo data and deductively with codes adapted from the World Vision Women’s Economic Empowerment (WEE) Framework and the UN Women’s Economic Engagement Framework (34, 35).

Data was then coded using the codebook in MAXQDA 2020 (VERBI Software, 2020). The analysis team comprised five team members (4 graduate research assistants [GRAs], each one responsible for coding data from one country), and a staff member (who read and coded all data to ensure codes were applied consistently and to gain insights across all countries). To ensure all who were coding had the same understanding of the codebook and how to apply the codes, a calibration activity was conducted among all team members engaged in analysis, wherein they independently coded the same transcript and met to discuss how codes were used and applied throughout the transcript. If there was disagreement, all discussed to reach consensus on how to apply the code going forward. The analysis team met weekly to discuss potential iterations to the codebook as needed.

Categories were developed from coded data to capture water-related enablers and barriers to women’s economic engagement. Furthermore, we identified patterns and linkages within and across the identified categories and organized them into an inductive conceptual framework to show how water impacted women’s economic engagement. The analysis team continually reviewed the data to ensure that the linkages and patterns were drawn from the data. By meeting weekly and discussing the data, the analysis team also was able to critically examine and reflect on any of their own biases and perspectives. Alongside having all data double coded (by one GRA per country and the supervising staff member across all countries), the weekly meetings aimed to reduce subjectivity in the analysis. Finally, two team members—who were not involved in coding but who brought extensive qualitative research experience and a broader knowledge of WASH and empowerment—provided input to assist in solidifying the framework.

### 2.6 Ethics and informed consent

The study was considered exempt by the Institutional Review Board committee of Emory University, Atlanta, Georgia, USA (IRB00005955), and was approved by the Comité de Ética del Centro Universitario de Occidente, de la Universidad de San Carlos de Guatemala (Acta 1.23 C.E. DICUNOC) in Guatemala; Universidad Nacional Autónoma de Honduras (CEIFCS-2023-P18) in Honduras; St. Paul’s University-Institutional Scientific Ethics Committee (ERB No. 38), the National Commission for Science, Technology and Innovation (NACOSTI/P/23/27117) in Kenya; and the Medical Research Council of Zimbabwe (MRCZ/A/3054) in Zimbabwe. All participants provided informed consent and participated voluntarily.

## 3.0 Results

### 3.1 Participants’ economic engagement experiences and water access

In all countries, women participants engaged in income-generating activities, though the proportion engaged varied by country. Specifically, 61% of women participants in Zimbabwe reported being engaged compared to 56% in Guatemala, 42% in Honduras, and 35% in Kenya. The types of income-generating activities women participated in were also variable, and included owning small businesses like selling second-hand clothes, accessories, and makeup; running grocery and food kiosks; keeping livestock like chicken, pigs, goats, cattle; selling animal products like eggs, milk, honey and products from smallholder farming like vegetables, tomatoes, and chilies. Some women created and sold products made from knitting, sewing (Guatemala, Honduras), weaving, bead work (Kenya), pottery and brick molding (Zimbabwe). Other paid work included doing laundry (Guatemala and Honduras); working in factories and harvesting coffee (Honduras), collecting pebbles, selling local brew, and harvesting and selling aloe and gum (Kenya); gold panning (Zimbabwe); and selling water and firewood (Guatemala, Zimbabwe). See Table 2 for more information on demographic characteristics.

**Table 2.** Demographic and water characteristics of focus group discussion participants, by country.

| Description | Guatemala<br>(7 communities) |  | Honduras<br>(6 communities) |  | Kenya<br>(6 communities) |  | Zimbabwe<br>(6 communities) |  |
| --- | --- | --- | --- | --- | --- | --- | --- | --- |
|  | Women | Men | Women | Men | Women | Men | Women | Men |
| **# Participants** | 54 | 34 | 64 | 39 | 80 | 80 | 100 | 49 |
| **Mean Age (SD)** | 38.42<br>(12.56) | 38.14<br>(12.56) | 40.41<br>(14.1) | 40.72<br>(14.71) | 42.41<br>(15.09) | 42.3<br>(14.98) | 46.00<br>(17.86) | 46.38<br>(18.33) |
| **Mean household size (SD)** | 6.3 (2.29) | 6.3<br>(2.29) | 5.2<br>(2.36) | 5.1 (2.26) | 6.4<br>(2.53) | 6.5<br>(2.55) | 5.5<br>(2.34) | 5.9<br>(2.43) |

| **Marital status, n (%)<sup>1</sup>** |  |  |  |  |  |  |  |  |
| --- | --- | --- | --- | --- | --- | --- | --- | --- |
| Single, never married | 8 (15) | 3 (9) | 6 (9) | 3 (8) | 1 (1) | 0 (0) | 9 (9) | 3 (6) |
| Unmarried, with partner | 17 (31) | 3 (9) | 39 (61) | 22 (56) | 2 (2.5) | 0 (0) | 0 (0) | 2 (4) |
| Married | 27 (50) | 28 (82) | 14 (22) | 13 (33) | 43 (54) | 80 (100) | 63 (63) | 42 (86) |
| Separated | 1 (2) | 0 (0) | 3 (5) | 1 (3) | 2 (2.5) | 0 (0) | 2 (2) | 0 (0) |
| Divorced | 0 (0) | 0 (0) | 0 (0) | 0 (0) | 0 (0) | 0 (0) | 3 (3) | 0 (0) |
| Widowed | 1 (2) | 0 (0) | 2 (3) | 0 (0) | 32 (40) | 0 (0) | 23 (23) | 1 (2) |
| **Completed schooling, n (%)<sup>2</sup>** |  |  |  |  |  |  |  |  |
| Never attended school | 11 (20) | 10 (29) | 9 (14) | 8 (21) | 75 (94) | 64 (80) | 9 (9) | 0 (0) |
| Some primary | 21 (39) | 13 (38) | 31 (48) | 15 (38) | 2 (3) | 10 (13) | 19 (19) | 11 (22) |
| Primary | 13 (24) | 5 (15) | 17 (27) | 12 (31) | 1 (1) | 1 (1) | 41 (41) | 20 (41) |
| Secondary | 7 (13) | 5 (15) | 3 (6) | 1 (3) | 1 (1) | 3 (4) | 31 (31) | 17 (35) |
| Above secondary | 2 (4) | 8 (26) | 1 (2) | 3 (8) | 0 (0) | 1 (1) | 0 (0) | 0 (0) |
| **Engaged in economic activities, n (%)** |  |  |  |  |  |  |  |  |
|  | 30 (56) | 26 (76) | 27 (42) | 35 (90) | 28 (35) | 35 (44) | 61 (61) | 22 (45) |

<sup>1</sup> *Marital status*: 1 missing men (Zimbabwe)
<sup>2</sup> *Completed schooling*: 3 missing men (Honduras), 1 missing men (Kenya), 1 missing men (Zimbabwe)

In all the countries, women were the primary persons responsible for water collection. Most women in Guatemala and Honduras had access to piped water, located in the dwelling or in the yard. In Kenya and Zimbabwe, most women collected water from unprotected springs and boreholes respectively, which were located somewhere outside their yard or plot. There was a slight variation in the mean water collection time per trip that women reported for drinking and other uses (including going to the water source, activities at the source, and returning). Women in Kenya reported the longest mean time for collecting drinking water (314 minutes), while those in Guatemala and Honduras reported the shortest time (19 and 25 minutes). Similarly, for water collection used for other purposes, women in Kenya had the longest mean time (326 minutes), and those in Guatemala had the shortest mean time (11 minutes). See Table 3 for more information on water-related characteristics.

**Table 3.**
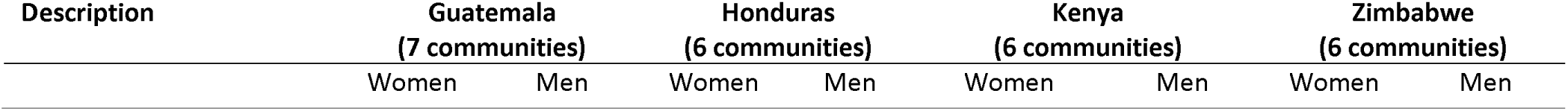
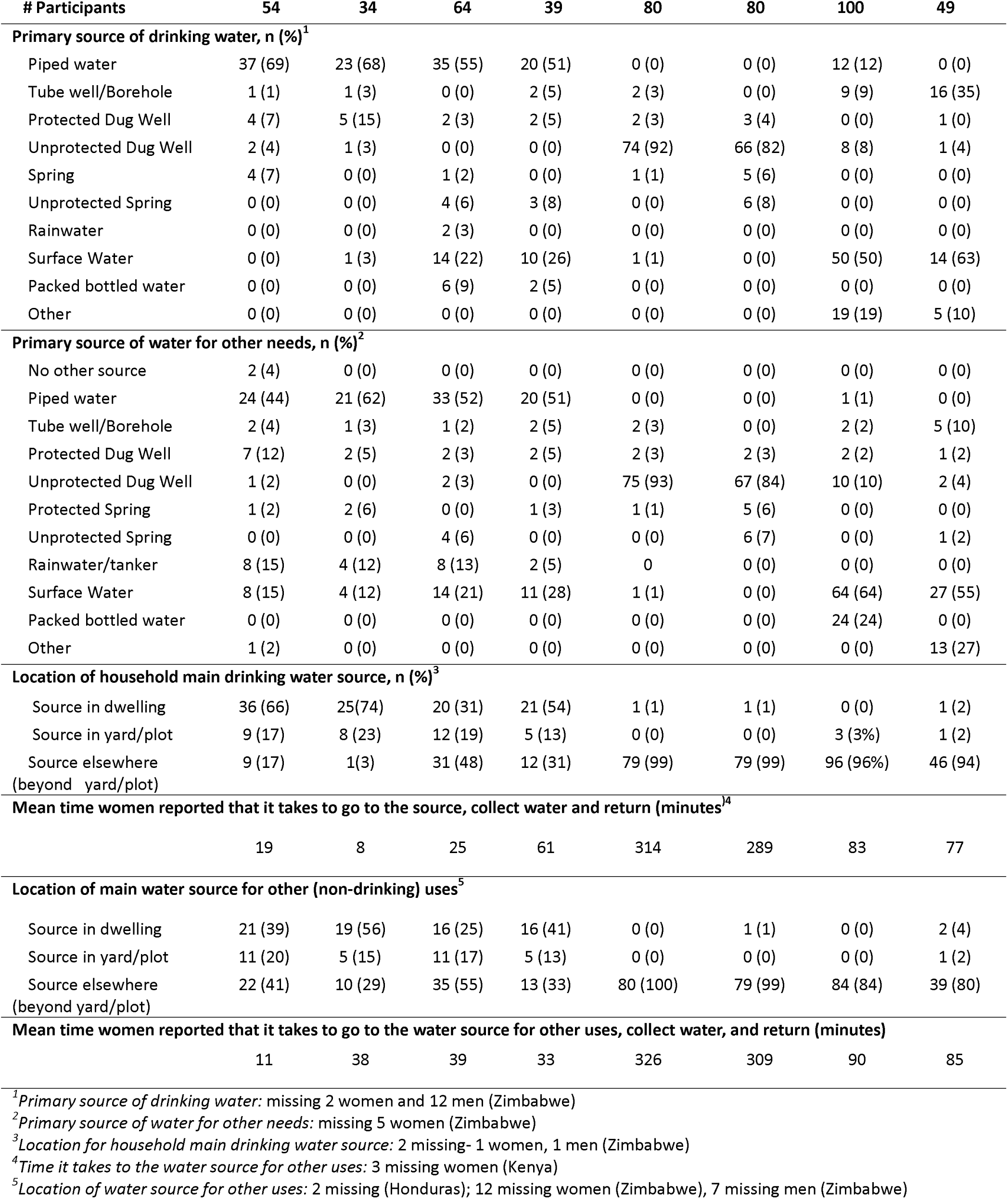
Water characteristics of focus group discussion participants, by country.

### 3.2 Relationship between water and women’s economic engagement

Based on participants’ insights, women’s economic engagement is influenced by their sufficient access to water. Specifically, access to water influences factors at the individual level and individual-level factors also influence water access. Further, environmental and societal level factors influence water access and individual-level factors (See Figure 1). Below, we first present and describe our inductive conceptual framework that shows how water and factors at other levels of social ecology (Individual, environmental, societal) influence women’s economic engagement. Next, we discuss the key domains within the conceptual framework in this order: individual level factors and impact on women’s economic engagement; environmental level factors that exacerbate water-related challenges and limit economic engagement; and finally, societal-related factors which cement the water-related challenges that constrain women’s economic engagement.

**Figure 1.**
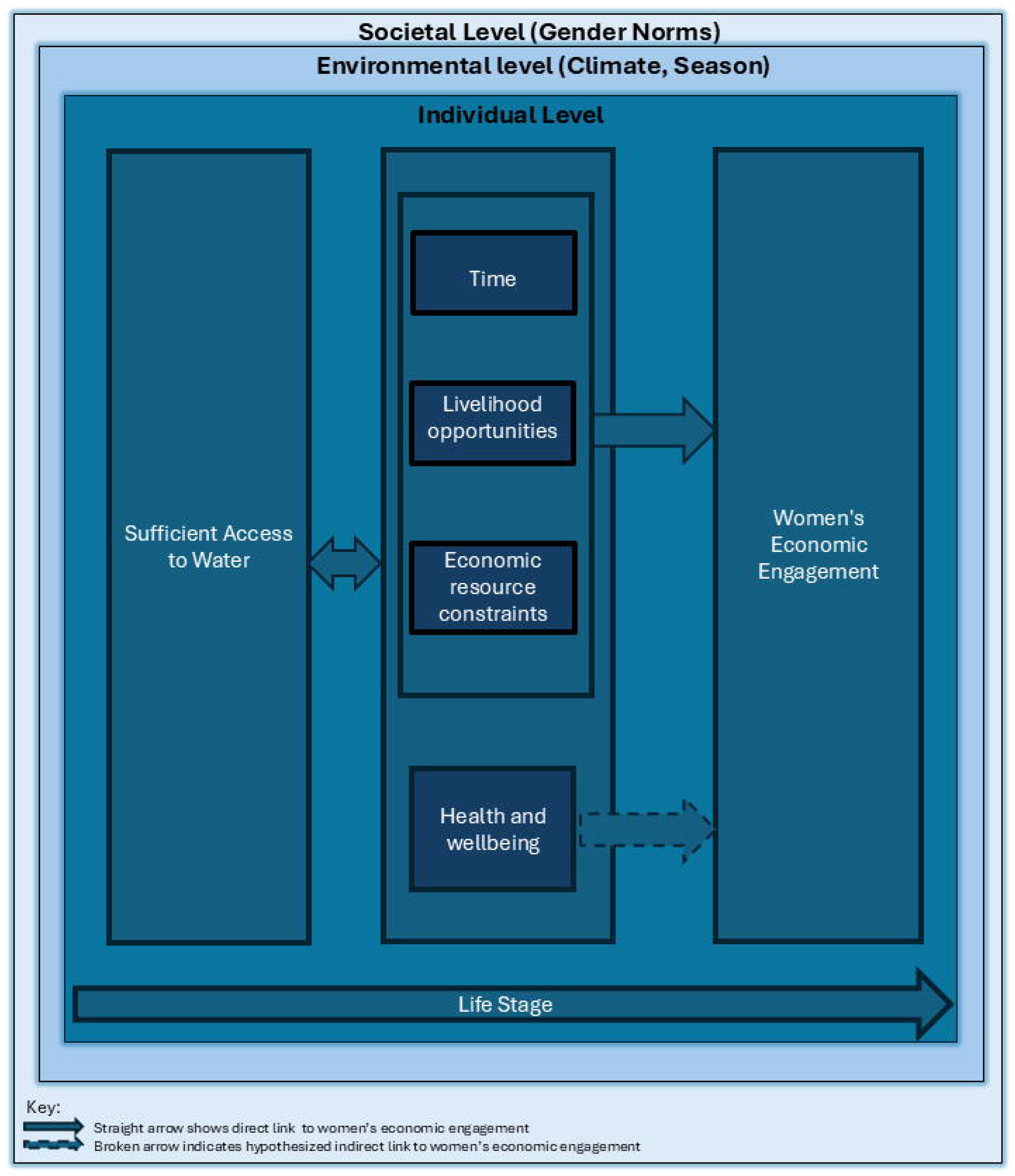
Conceptual Framework on Water and Women’s Economic Engagement.

Based on participants’ reflections, women’s economic engagement is influenced by factors at the individual, environmental and societal levels (Figure 1).

The central components of the conceptual framework (Figure 1) show the relationship between sufficient water access, individual level factors—including time, livelihood opportunities, economic resource options, and health and wellbeing—and women’s economic engagement. Each of the individual-level factors will be discussed in depth below. In brief, when women do not have time, which can be compromised by limited access to sufficient quantities of water, they cannot engage in economic opportunities. Livelihood activities like agriculture and livestock production that women engaged in can be reduced due to lack of access to water. Women’s economic resources can be constrained due to water, for example if they need to pay for water or for costs associated with water access, which can limit the economic resources they may need to participate in income-generating activities. Lastly, health and wellbeing, specifically, physical health issues like fatigue, exhaustion, physical injuries, and pain, and from water collection work, can leave women depleted of energy needed for economic engagement activities.

Our conceptual framework further shows that societal, environmental and life stage factors exacerbate the influences of access to water on individual level factors and women’s economic engagement (Figure 1). Gender norms shape roles and responsibilities for men and women and ascribe water collection as the primary responsibility of women and girls in all four countries. Environmental factors, such as drought and seasonality, diminished sufficient access to water, further reducing women’s time and energy as they had to go to more distant sources, and negatively impacted livestock and agricultural productivity due to diminished pasture and reduced water supply.

### 3.3 Individual level factors and Impact on Women’s Economic Engagement

Women’s ability or lack of ability to access water in sufficient quantities influenced both their lives and the options available for their economic engagement. Water collection is among one of the many tasks women are expected to do, and is a priority task because water is necessary both for survival and to complete many of their other responsibilities. Further, the availability of water dictated how they pursued activities beyond their responsibilities, including rest. Participants in Kenya and Guatemala summarized the primacy of water in their lives:

> “Water is a basic need. As a woman you can’t be able to have your things alright and arranged when there is unavailability of water in the household. We get a lot of stress when we lack water, you can’t do anything without water…” FGD2(W): Community 1, Kenya

> “There is no life without water; nothing will survive without water either for the animals or people.” FGD2(M): Community 5, Kenya

> “A woman’s life is really hard. We must take care of the children, wash clothes, clean and carry out all the housework. We go to bed late. We don’t have time to sit down and rest. We do other jobs. We get up at three in the morning…We get up early to do our tasks. That’s what we do. At the moment, what we need the most is water.” FGD1(W): Community 5, Guatemala

Women reported that lack of sufficient water was a crucial factor that hindered them from engaging in economic activities. Specifically, they discussed how sufficient access to water impacted their (a) available time, (b) livelihood opportunities, (c) access to economic resources, and (d) health and well-being, all of which influenced their ability to engage in economic activities. Each of these is discussed in turn below.

#### 3.3.1 Time

Lack of access to sufficient water and consequent related water-work influenced the time women had available to engage in economic and other activities. In this context, time refers to the amount of time needed for water collection and other water-related work, as well as when in the day women can participate in economic activities.

How women spent their time each day largely was determined by water, including the quantity and quality of water needed, the distance to and availability at the source, and the water-related work they needed to do. For many women, the water collection process could take an extensive amount of time, given the great distances women had to walk to get to water sources and back. Their long journeys to the sources could be further slowed if they faced rough terrain, needed to prioritize livestock or give way to wild animals at a water source (e.g., elephants in Kenya), or carried heavy weights (e.g., water, children, laundry). In Kenya, participants reported that it could take as many as eight hours to travel up to 20 kilometers on a round trip to get water, and in Zimbabwe, participants said it took between two and five hours to collect water, and potentially longer in times of drought or during dry seasons. Women in Guatemala, Honduras, and Zimbabwe discussed having to make multiple trips per day to the water source when water was available at the source, leaving them with little time to complete their other household duties, let alone engage in income-generating activities. And women in Kenya expressed that most of their day was devoted to water collection, as described by one man:

> “We have many challenges, but the greatest of all is water, we have water problem in this area which is the main reason why many women cannot engage in economic activities. The women will take a donkey to the river, and they will use the whole day from morning to the evening because the water source is very far from the village.” FGD(M): Community 5, Kenya

Beyond the time needed for going to and from the source, women from all countries reported that they required substantial amounts of time at the source, further compromising their ability to plan for and attend to other activities, including meetings related to economic activities in the community, economic ventures, and even household responsibilities. At water sources, women spent time waiting in queues, doing laundry, pumping at rusty or old boreholes, scooping water from dry riverbeds, or searching for clean water, which also diminished their energy. Due to the extensive amount of time and energy for this water related work, women were exhausted, and prioritized completion of household chores before economic activities. In all countries, women had a clear hierarchy of activities to perform, with the expectation that household duties were completed before economic activities could begin. Women still faced time-burden challenges even where water sources were close to their homes, specifically in Guatemala, Honduras, and some communities in Zimbabwe. Women reported experiencing increased waiting time for water due to absence of water operators at the source (Zimbabwe), intermittent water supplies due to water rationing (Guatemala, Zimbabwe), breakages of pipes from heat (Guatemala), clogging of pipes during heavy rains (Honduras), and insufficient and delayed maintenance of pipes, which constrained access (Guatemala, Honduras, Zimbabwe). As a result, women said they opted for alternative water sources which were further or waited longer to get water, which diminished their time. See Table 3 for illustrative quotes related to time.

**Table 3.** Illustrative quotes about time, by country.

| Guatemala | Honduras | Kenya | Zimbabwe |
| --- | --- | --- | --- |
| "My life would change significantly. I would have more time to devote to my children and my family. You could even look for a job. Sometimes when a job opportunity comes up, I must turn it down because I must go to the river, that's something I always bring up. If I had access to water in my home, I ... it would greatly change my life." FGD(W): Com1<br>"In addition, this would save them time, since if there was water, they could carry out washing tasks at any time. Sometimes, some women express that, even though they go out to work, they would not have to set aside specific time to attend to issues related to water if it were available on many occasions. If there was water, sometimes we would wash a few clothes in the afternoon, when we get up in the morning to wash." FGD(M): Com5<br>"What prevents us from participating in economic activities is the lack of time. I could be working to generate income. However, I find it difficult to meet the deadlines required for the work assigned to me, which in this case is yarn production. The reason is that I waste three to four hours a day going to the river to get water, and I would need that time to dedicate myself to my work and save some money. If I had access to clean water at home, I could spend that time working on my yarn..." FGD(W): Com1 | "Certainly, having access to water at home would save time spent traveling to the river. This extra time could be used for various tasks like housework, caring for plants, and irrigation. Currently, I don't have help at home, so when I return from fetching water, I often need to prepare food even when tired and hungry. Having water at home would make household tasks easier." FGD(W): Com2<br>"The time they spend bathing or collecting water from the stream, at least, would be used to prepare donuts that they can sell, with the purpose of acquiring at least a pound of sugar. This is because the stream is located far away." FGD(M): Com1<br>"Now, instead of wasting time going to the river, we no longer waste it transporting horses, since we have water in our own house. This allows us to use that time on other activities that we used to miss by having to go get water from the river or stream." FGD(W): Com4 | "I can do some small business here, but I don't know how to do it without water around, people normally stay even two days without water and yet how do business work if there is no water, it won't work surely. Sometimes we stayed without drinking tea leave alone having eaten food for some days, so business won't work here just because of the water's unavailability." FGD(W2): Com5<br>"Life will change when you have water at home because you will have time to engage in other activities which generate income and also, the young children and old people will not have problems again so their health will be better and even the livestock will not be suffering anymore." FGD(M1): Com6<br>"Time is not enough to engage in economic activities because where we go, and fetch water is very far. Sometimes you can ask your child not to go to school that day, but you cannot ask the child not to go to school two times, so water is a major problem that is hindering us from engaging in economic activities." FGD(W2): Com2<br>"A woman that wants to do businesses when will she do the business, and she goes to the river early morning and comes back at 4pm what kind of development is this person doing that comes in the evening with little water and in the morning, she has to go back there?" FGD(M1): Com4<br>"What I want to say is the water challenge have made us not to engage in other developments, our youths or women tried to engage in business activities, but they fail due to water problem because most of their time is used to search for water." FGD(M2): Com5 | "Time depends on where you stay and whether you get to the water source with no queue. To go to the tap, you need to travel three times because you will need to carry 3 buckets, so you need a lot of time. The borehole is nearer, but the water is rustic and if you go alone you need to wait till you find someone to assist you to pump the borehole because it's hard and heavy to pump, it [the pump] is now old." FGD(W1): Com3<br>"We carry out a lot of economic activities, but we do not do it well because of water crises and scheduling. If water is closer to my homestead, I have ample time to plan for other activities unlike now where we spend time ferrying water. Our business does not operate well because of water; I end up opening my business at 10 am after fetching water." FGD(W2): Com1<br>"We have other activities like sewing but we do not have time to do it since we spend more time doing water journeys." FGD(W2): Com2<br>"My challenge is the distance. It disrupts my plans for the day because I must walk long distances. It has made me schedule days to fetch bulk quantities of water so that I have time for other things" FGD(W1): Com1<br>"We live close to the borehole, but it takes us more time to pump the water. We wait for about 2hours for the water to start coming out and it will be dirty. When it gets a bit clearer that's when we pump into our buckets." FGD(W3): Com2 |

In some communities in Guatemala, water -work included mandatory service on water boards and committees for every household for one to two years, which added substantial burden to the already limited time that women could utilize for economic activities. The time women spent serving on water committees diverted their time away from economic-related activities like weekly markets, produce production and sales, and participation in training and loan group meetings, and made it challenging to balance household responsibilities. This mandatory service disproportionately affected single and widowed women as women reported that they had to shoulder the responsibility since they did not have partners to share the responsibility.

> “It happens that many of the committee members are single mothers or widows. If the service is in their name, then they have to assume their position on the committee when appropriate. This is why this year the committee is made up mostly of women, since it has been their turn to assume that responsibility.” KII(F): Community 3, Guatemala

In all countries, participants indicated that consistent access to sufficient and quality water could alleviate the time burden associated with water collection and water work, allowing them to dedicate the time saved to other activities. Specifically, women mentioned that they would do housework, childcare, rest, sleep longer, and engage in income-generating activities including livestock-rearing (poultry, pig, cattle, and goats) and crop production. In Honduras, one woman mentioned that she would do other activities like household activities quickly to have time to engage in economic activities and another said the time would be used to take care of grandchildren and not necessarily for engaging in economic activities. In Guatemala, women mentioned that they would use the time to work, care for children, wash clothes, and rest. In Kenya and Zimbabwe, women indicated that they would sleep longer, rest, and engage in gardening and income-generating activities.

> “I would engage in poultry farming and avoid the long journeys to fetch water. It would give us more time to rest our old bodies and consider money-making projects like poultry, selling eggs, and making more money.” FGD(W): Community 2, Zimbabwe

The need to prioritize water collection for household needs over livelihood activities threatened the sustainability of economic activities women engaged in. Women reported that even if they had products to sell, water collection time dictated when they could sell products, negatively affecting the quality of their businesses. For example, in all four countries, participants sometimes had to open businesses or other income-generating activities late in the day or close their businesses to prioritize water collection. One woman from Kenya describes how water fetching impacted her business opportunities:

> “This water problem is also not letting us engage in businesses because if you have things that you wanted to sell but you have no water (at) home so you decide to go fetch water and while on the way some people come to look for goods while you are away, and they go back and get what they wanted from somewhere else. Has that not led to the fall of your business? Your business just falls due to that.” FGD(W): Community 5, Kenya

#### 3.3.2 Livelihood Opportunities

Women reported that livelihood activities -- defined here as income-generating activities that require water for agriculture, livestock production, pottery, brick molding, laundering clothes for money, and water vending -- were limited by insufficient water access, diminishing women’s economic opportunities. Women’s inability to access sufficient quantities of water hindered their ability to take part in businesses deemed profitable, like food production in Guatemala, pig-farming in Honduras, goat-rearing in Kenya, and poultry projects in Zimbabwe. Participants in Honduras, Kenya, and Zimbabwe reported that the lack of water led to animal disease, contributing to the thinning and death of livestock and influencing their ability to start up and sustain existing economic activities. Additionally, human-animal conflicts over water led to the loss of livestock, which in turn impacted their economic activities. See Table 4 for illustrative quotes related to livelihood.

**Table 4.**
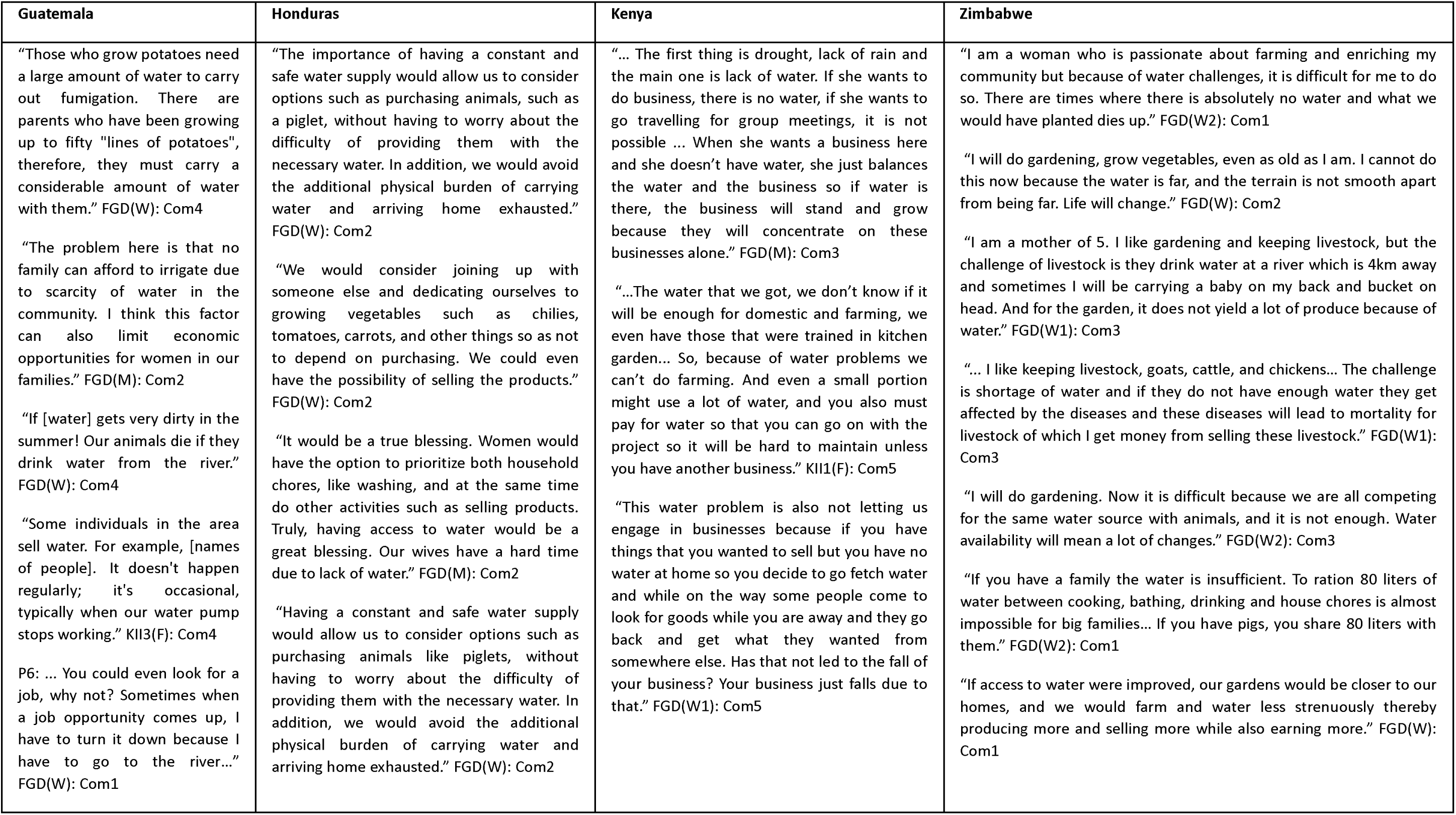
Illustrative quotes about livelihood, by country.

| Guatemala | Honduras | Kenya | Zimbabwe |
| --- | --- | --- | --- |
| “Those who grow potatoes need a large amount of water to carry out fumigation. There are parents who have been growing up to fifty “lines of potatoes”, therefore, they must carry a considerable amount of water with them.” FGD(W): Com4<br>“The problem here is that no family can afford to irrigate due to scarcity of water in the community. I think this factor can also limit economic opportunities for women in our families.” FGD(M): Com2<br>“If [water] gets very dirty in the summer! Our animals die if they drink water from the river.” FGD(W): Com4<br>“Some individuals in the area sell water. For example, [names of people]. It doesn't happen regularly; it's occasional, typically when our water pump stops working.” KII3(F): Com4<br>P6: ... You could even look for a job, why not? Sometimes when a job opportunity comes up, I have to turn it down because I have to go to the river...” FGD(W): Com1 | “The importance of having a constant and safe water supply would allow us to consider options such as purchasing animals, such as a piglet, without having to worry about the difficulty of providing them with the necessary water. In addition, we would avoid the additional physical burden of carrying water and arriving home exhausted.” FGD(W): Com2<br>“We would consider joining up with someone else and dedicating ourselves to growing vegetables such as chilies, tomatoes, carrots, and other things so as not to depend on purchasing. We could even have the possibility of selling the products.” FGD(W): Com2<br>“It would be a true blessing. Women would have the option to prioritize both household chores, like washing, and at the same time do other activities such as selling products. Truly, having access to water would be a great blessing. Our wives have a hard time due to lack of water.” FGD(M): Com2<br>“Having a constant and safe water supply would allow us to consider options such as purchasing animals like piglets, without having to worry about the difficulty of providing them with the necessary water. In addition, we would avoid the additional physical burden of carrying water and arriving home exhausted.” FGD(W): Com2 | “... The first thing is drought, lack of rain and the main one is lack of water. If she wants to do business, there is no water, if she wants to go travelling for group meetings, it is not possible ... When she wants a business here and she doesn't have water, she just balances the water and the business so if water is there, the business will stand and grow because they will concentrate on these businesses alone.” FGD(M): Com3<br>“...The water that we got, we don't know if it will be enough for domestic and farming, we even have those that were trained in kitchen garden... So, because of water problems we can't do farming. And even a small portion might use a lot of water, and you also must pay for water so that you can go on with the project so it will be hard to maintain unless you have another business.” KII1(F): Com5<br>“This water problem is also not letting us engage in businesses because if you have things that you wanted to sell but you have no water at home so you decide to go fetch water and while on the way some people come to look for goods while you are away and they go back and get what they wanted from somewhere else. Has that not led to the fall of your business? Your business just falls due to that.” FGD(W1): Com5 | “I am a woman who is passionate about farming and enriching my community but because of water challenges, it is difficult for me to do so. There are times where there is absolutely no water and what we would have planted dies up.” FGD(W2): Com1<br>“I will do gardening, grow vegetables, even as old as I am. I cannot do this now because the water is far, and the terrain is not smooth apart from being far. Life will change.” FGD(W): Com2<br>“I am a mother of 5. I like gardening and keeping livestock, but the challenge of livestock is they drink water at a river which is 4km away and sometimes I will be carrying a baby on my back and bucket on head. And for the garden, it does not yield a lot of produce because of water.” FGD(W1): Com3<br>“... I like keeping livestock, goats, cattle, and chickens... The challenge is shortage of water and if they do not have enough water they get affected by the diseases and these diseases will lead to mortality for livestock of which I get money from selling these livestock.” FGD(W1): Com3<br>“I will do gardening. Now it is difficult because we are all competing for the same water source with animals, and it is not enough. Water availability will mean a lot of changes.” FGD(W2): Com3<br>“If you have a family the water is insufficient. To ration 80 liters of water between cooking, bathing, drinking and house chores is almost impossible for big families... If you have pigs, you share 80 liters with them.” FGD(W2): Com1<br>“If access to water were improved, our gardens would be closer to our homes, and we would farm and water less strenuously thereby producing more and selling more while also earning more.” FGD(W): Com1 |

Flourishing agriculture and large-scale crop production for economic gain are dependent on water availability and are diminished when insufficient quantities of water are available. In Honduras, Guatemala, and Zimbabwe, water shortages were linked to women’s inability to grow vegetables— such as tomatoes, butternut squash, onions, chilies, and leafy greens—for consumption and sale. In Kenya, women reported not being able to grow vegetables at all, even though they expressed wanting to garden. Despite their existing knowledge on irrigation, women reported not being able to do much in terms of agriculture because of lack of water.

Although participants in all four countries described the negative effects of insufficient water access, they also noted that access to water sources close to their homes that provided sufficient quantities of water could enable economic opportunities. As one woman from Zimbabwe reported, “If we have water, we have money.” Women described how such sources could ensure healthy and quality livestock, facilitate their ability to grow vegetables to sell, and enable other water-dependent businesses, like clothes laundering. In Guatemala, one participant mentioned that if she had access to water at home, she would have the flexibility to carry out her activities without worrying about going to the river, and that improved availability and access to water could additionally allow women to grow crops and rear livestock, contributing to income generation. One woman from Zimbabwe described the critical role of water and how improved access would open economic opportunities. Women in all four countries reflected on the potential for economic opportunities if water was available:

> “In many ways, if there was water, we could have orchards. Who wouldn’t have money? Who wouldn’t have a business? Even if we didn’t have a place to plant, it is easier to rent a place to plant, but without water it is not possible.” FGD(M): Community 2, Guatemala

> “Sometimes there are neighbors who are looking for someone to wash their clothes. Having water in my house, I could take care of this job because I wouldn’t have to go wash it in the river. So, in my case, access to water would already be of great help to me.” FGD(W):

Community 4, Honduras

> “When water is near, it will help us a lot because there are some areas we border, and we have seen how they work. If we get water, people will start other ways of survival. They will cultivate and grow things like vegetables. Areas like [name of a place] have water and they have grown things like kales and other vegetables and those villages are benefiting a lot.” FGD(M):

Community 2, Kenya

> “At times it is lack of water. Water sources are far for women to get water to start garden projects and to grow vegetables to sell so that they can get capital. We go far to get water from Silikwe River and at times from Umzingwane River. If there is water nearby, we can do garden projects and get capital. Water is the most crucial factor that hinders us from starting businesses.” FGD(W): Community 2, Zimbabwe

#### 3.3.3 Economic resource constraints

Economic resource constraints, specifically, direct and indirect costs of water, limited the capital women had to enable their participation in income-generating activities. Some women paid for water directly, while others reported costs associated with water access, such as pipes, transport, water storage vessels, and operations and maintenance of the water sources. These costs varied by household and by community. For instance, in Guatemala, participants shared that people may pay up to $35-40 per 1,000 liters to those who fetch water using their cars. Additionally, they also reported that women from “richer households” (perhaps those that have a household borehole, well, or a car) sell water to other community members for between $20-30 per 1,000 liters. In Honduras, a small bag of water would cost approximately $1, whereas in Kenya, a 20-liter container of water would cost $2 which cumulatively would add up to $60 per month for water expenditure. With families already struggling with poverty due to loss of livestock--the sole source of household income--women were forced to use their limited funds to pay for water, which would not be enough to sustain farming for income generation.

> “So, because of water problems we can’t do farming. And even a small portion might use a lot of water, and you also must pay for water so that you can go on with the project so it will be hard to maintain unless you have another business.” KII(F): Community 5, Kenya

#### 3.3.4 Health and wellbeing

Women’s physical health and mental wellbeing were impacted by a lack of access to water, further limiting their ability to engage in other activities. While women did not directly state that their health and well-being was impacted by water and impacted their ability to engage in economic activities, many women reported experiencing fatigue, exhaustion, physical injuries, and pain from water collection work, which left some so depleted that they said they had no energy left to do anything else. One man from Zimbabwe remarked on how water work was so demanding it does not allow women the ability to think about a business.

> “The issue of water availability is a serious challenge. I am sure you haven’t met any voluptuous woman since you came into our community. Our women work too hard to even accumulate any fat. They are always very, very slim. There is no time and energy left to think of business after the suffering and struggling [for water].” FGD(M): Community 2, Zimbabwe

See Table 5 for other illustrative quotes related to health and wellbeing.

**Table 5.**
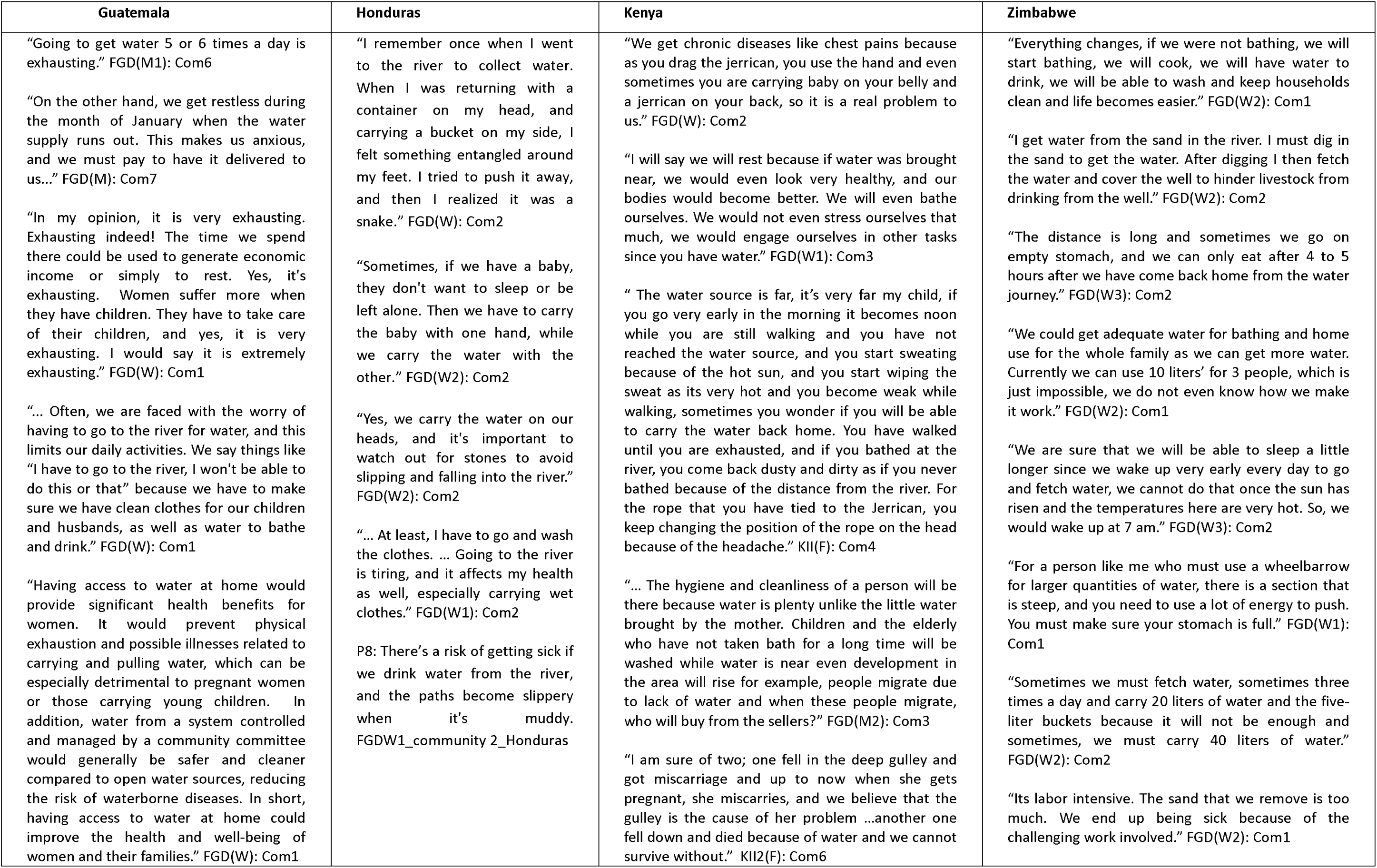
Illustrative quotes about health and wellbeing, by country.

Beyond walking long distances and carrying heavy weights, water collection could also involve strenuous processes that strained women’s energy. In Guatemala and Honduras, women filled and hauled large containers with water when they had access because flow was intermittent, and they wanted to ensure that they had enough water supply to last when water was not flowing. Moreover, due to inconsistent access, women sometimes had to make multiple trips to ensure they could get water when it was available. In Zimbabwe, some women pumped water from rusty pumps, requiring much energy. In Kenya and Zimbabwe, women engaged in sand abstraction which required them to dig at dry riverbeds and scoop out sand until water started coming out, then wait for the water to clear up and use a small bowl or container to collect and pour the water into jerricans to carry home. To get this water home, some women in Zimbabwe and Kenya pushed heavy wheelbarrows and scotch carts holding their water containers or rolled their heavy jerricans on the ground if they lacked scotch carts or donkeys. This form of hauling water causes strain on women, especially elderly, sick or pregnant women.

In all the countries, women reported risks of physical injuries along their journeys to reach a water source, as well as physical pain from hauling water. Injuries could result from the steep and rough terrain or even encounters with wild animals, including elephants. In Kenya, participants reported that other women in the communities experienced miscarriages or broken bones from the challenging walk to the water sources. Despite pain involved with water collection, women had to persist as best they could. One woman in Honduras described how she limited the amount of water she collected due to pain she experiences carrying water with an existing back problem:

> “In my situation, when I go to pick up my bucket of water, I only make two trips because afterwards I experience pain in my spine and find it difficult to get up. I have back problems, so carrying the bucket from where I go to get water twice affects me. Therefore, I need someone to help me lower my bucket of water when I go with someone else, since my spine no longer allows me to do it on my own. Sometimes, I find myself having to ask a child to accompany me and help me.” FGD(W): Community 4, Honduras

Women also reported being stressed by thinking about water collection processes and related activities. Women reported thinking about where to get water, queuing for water, the distance to the sources, childcare during water collection, and resultant effects of lack of water in the households, like children not being able to eat or attend school. One woman in Kenya described this mental burden:

> “Even when you are sleeping, you start planning for your things as you sleep like you start saying tomorrow, I do not have water. You pray even for God to give you that water early. When you wake up in the morning you already have plans for the entire day. After I collect water, I need to clean the clothes, I need to go somewhere; I need to be cooking, so, all those are woman tasks.” FGD(W): Community 4, Kenya

### 3.4 Environmental-level factors exacerbate water-related challenges constraining economic engagement

Environmental factors, like seasonality and drought, were reported to impact water sources, exacerbating water-related challenges that constrain women’s participation in economic activities. Across the four countries, participants reported that water levels had noticeably reduced in dams, rivers, and groundwater tables due to drought and seasonality, reducing the quantity of water participants would have access to for meeting household and other needs, like cooking, drinking, personal hygiene, livestock and agricultural production, and economic opportunities that require water. Across all the countries, participants continually noted that many of their struggles were either caused by or worsened by a lack of water. In Kenya and Zimbabwe, prolonged drought vastly decreased water availability at the water sources, with adverse effects on agriculture and livestock. In Honduras and Guatemala, participants reported seasonality affecting water availability, as water scarcity was more acute in summer, during the dry season. The decrease in water availability due to seasonality or drought necessitated women to use alternative water sources like springs, wells, and dams, which participants reported to be farther from their homes. These alternative sources provided insufficient quantities of water, were shared by domestic and wild animals, and were not safe for consumption. Additionally, geographical factors also exacerbated water scarcity, as some communities were in places where there were no natural water sources or had low groundwater potential which could not allow for drilling boreholes and wells.

Life stage lessened women’s engagement in economic opportunities. The burdens exacerbated by seasonality and drought disproportionately affected pregnant women, women with young children, and elderly women. For example, in Honduras, Kenya, and Zimbabwe, elderly people, pregnant women and women with young children reported strain from walking long distances to collect water from alternative water sources. Older and pregnant women have greater challenges walking long distances to the water sources, carrying water due to the weight burden, and making multiple trips. Women with young children may need to carry them to the water source, which increased weight burden and depleted energy and time. These experiences minimize women’s livelihood options by reducing their time and affecting their health and wellbeing. (See illustrative quotes describing life-stage related experiences in Table 6.)

**Table 6.** Illustrative quotes about life stage, by country.

| Guatemala | Honduras | Kenya | Zimbabwe |
| --- | --- | --- | --- |
| “It happens that many of the committee members are single mothers or widows. If the service is in their name, then they have to assume their position on the committee when appropriate. This is why this year the committee is made up mostly of women, since it has been their turn to assume that responsibility.”<br>KII3(F): Com3<br>“Single mothers, particularly those without husbands, may request their own water sources to the committee.”<br>KII2(F): Com3 | “In the case of single mothers, they have sole responsibility for making these decisions [utilization of resources] for themselves and the children.” KII1(M): Com4<br>“Of course, there are difficulties for women. Some have just given birth or have babies that are months or even days old. These women cannot go to collect water... In such a trip, they can only take their babies with them. They would not be able to carry water. They can't even carry diapers, or much of a bucket of water. FGD(M): Com1<br>“One's age would be one obstacle. The elderly no longer possess the strength to retrieve water.” FGD(M): Com2 | “... even a pregnant woman must go and drag the jerricans from water and by doing so, she might even start to bleed and even miscarry. Even the one who gave birth she has nobody to bring water for her ...”<br>FGD(W): Com3<br>“It is just like that, you wake up and go to collect water even if you are old, and you go and meet with elephants here but you cannot come back home so you watch them where they are going so that you pass the other side, so you stay there until when God gives you somewhere that you can pass, so you go and collect water and come back taking rests under trees because of the hotness of the sun.”<br>FGD(W): Com 5 | “It is too far. For elderly people it is not good at all to walk such long distances.”<br>FGD(W): Com2<br>“Elderly women like that grandmother of ours face difficulties in fetching water because of distance and amount of water carried. They cannot go two or three times a day, they can spend the whole day moving up and down to fetch water and they use small containers because they cannot carry big loads at once because of the amount of energy used. They will not be able to do anything else.” FGD(W): Com3<br>“The water source for me is not easy to access, since I cannot walk long distance due to old age. At the same time when I manage to go, I can only carry one bucket which has to do my dishes, bath, cook and drink as well.” FGD(W): Com2 |

### 3.5 Societal-level factors cement water-related challenges constraining economic engagement

In all countries, women’s ability to engage in income-generating activities was constrained by gender norms requiring them to handle all domestic labor, including the provision of water. Gender norms determined women’s and men’s household roles and responsibilities. In all the countries, men, as household heads, dictated what women can and cannot do and women were largely expected to take care of all unpaid household duties, like fetching water and firewood, childcare, cooking, cleaning, laundry, and attending to small animals, even if they were engaged in other paid work. In Honduras, for example, men and women perceived women as keepers of homes, and women were described as housewives whose role was in the kitchen; they were expected to take care of household chores in addition to taking care of their children and husbands. In Guatemala one man indicated that “los hombres son superiores” (men are superior), and that women themselves did not have a voice and would not be involved in leadership. Furthermore, some women in all countries indicated that they had to seek permission from their partners to be able to engage in income-generating activities as well as how money was spent. See Table 6 for illustrative quotes related to gender norms.

**Table 7.** Illustrative quotes about gender norms and water, by country.

| Guatemala | Honduras | Kenya | Zimbabwe |
| --- | --- | --- | --- |
| "It happens that many of the [water] committee members are single mothers or widows. If the service is in their name, then they have to assume their position on the committee when appropriate. This is why this year the committee is made up mostly of women, since it has been their turn to assume that responsibility." KII2(F): Com3<br>"Women, like me, take care of this [water collection]. The men go out to work and come back tired, while we [women] take care of the water and everything at home, since we [women] are the housekeepers." KII1(F): Com6 | "This happens [women collect water] because the men work in the field and, sometimes, are not available. However, when they can't do it [collect water], we go and do it ourselves. The main reason why women are mostly responsible for fetching water is that most men, such as myself, spend very little time at home due to our work in the fields and our long days in the mountains." KII1(M): Com 2<br>"For the most part, women are in charge of collecting water in this community, often bringing their children with them to help with the task... This task [water collection] is traditionally associated with their responsibilities, since they are housewives and in charge of the kitchen." KII2(F): Com3<br>"From time to time, men go to collect water, but generally it is us women who go. This is because men are usually not at home, but if they realize that we need water, they go to the river. Although this is not daily." FGD(W): Com2 | "According to the Samburu culture it's role of a woman to fetch water in this community either you like or not it's a taboo for a real man to fetch in its community while his wife is still alive unless she has died that the only way that you might see a man going to collect water from anything else. ... When other men see another man helping his wife, he will be isolated by his fellow men for sure, and no one will entertain that man." FGD(W2): Com5<br>"Because here everybody has their roles, and roles for the husbands are to take care of the livestock and the wife's role is to take care of the house. So, if it's drinking water, it's the mother, livestock is for men so that's why the mother has no other role than that of the house." KII1(M): Com5<br>"According to Samburu culture it's shameful for a man to fetch water in Samburu community since men have their roles to play on in the community. So, women are the one who fetch water for household because they are the one who take care of their children. A man doesn't care if there's water or not their work is only to get food ready..." FGD(W): Com5 | "It's us women who go to collect water, men will say it is not possible for men to carry a bucket and go and collect the water." FGD(W1): Com1<br>"We ask them [men] to go with us so they can help but they just dismiss the issue because they feel it's women's duty to fetch water, so they do not know how difficult it is to pump water out of the borehole." FGD(W3): Com3<br>"Men do not want to fetch water. They say it is for women. If we go to the farm together when we come back, he will just come and rest under the tree shade and I, the women, will start washing the dishes and plates after that I will take the buckets and go to fetch water. They will be saying be quick we want to go back to the field." FGD(W1): Com3<br>"Women are mainly responsible for water collection. They are the ones solely responsible for these chores. Men from most households will be away in small scale mining and some working away from their homes. Water collection is mainly for girls and women. Men assist in rare cases." KII1(M): Com2<br>"Women are the main custodians of collecting water. They are responsible for the everyday collection of water. Man will collect water less frequently." FGD(M): Com2 |
| Illustrative quotes about general gender norms, by country |  |  |  |
| "... We are still rooted in the principles of past generations, where women had no voice or leadership. For example, there are no women in the Community Development Committees (COCODES), and this is because no one allows them to participate. Even mothers and fathers who have grown up with these beliefs still feel that fear, where men are considered superior. This mindset persists today." KII2(F): Com4<br>"In this community, it's a tradition for women to focus on homemaking while also taking care of their families. FGD(W): Com4 | "Yes, because some are sexist. On several occasions I have heard some men comment, 'Why are those women selling on the street? They should be at home taking care of their families.'" FGD(W): Com4<br>"At times, even when we are absent, is it necessary for the women to go and fetch water because running out of it is not an option." FGD(M): Com2 | "Most of the time men are the ones to decide as I have told you earlier, men have that ego that they are the head of the family, so they are the ones to make decisions." KII1(F): Com2<br>"For women may be for the single mothers and widows can just decide for themselves depending on but if you have a husband, you know mostly the husband is the one who makes." KII1(M): Com4 | "What I can say is my barrier is I do not have a man to assist me [with responsibilities]. I have too many responsibilities on my shoulders, and I cannot manage." FGD(W2): Com1<br>"I agree with what she is saying. In our culture the man is the head of the household hence you cannot decide outside the man." FGD(W2): Com2 |

Men and women in all countries reported that water collection was the primary responsibility of women and older girls, and occasionally younger children, but was not expected of men. Men and women noted that this responsibility particularly constrained women’s abilities to engage in paid economic activities. Across all countries, gender norms prevented men from engaging in water collection, except in Guatemala where women reported that men helped in times of scarcity. In Honduras, while women reported that water collection was a shared responsibility at the household, men would typically be busy working in the fields and were only able to collect water occasionally. In Kenya, women mentioned that it was shameful for men to collect water, and the men involved in water collection were perceived as “madai” [stupid] and faced isolation in the community. In Zimbabwe, cultural norms dissuaded men from water collection. Women indicated that men did not engage in water collection work, did not know how difficult pumping water from the borehole is, and perceived that it was impossible for men to carry buckets to go to collect water.

## 4.0 Discussion

Findings from this qualitative study to assess the role of water in women’s economic engagement in selected rural areas of Guatemala, Honduras, Kenya, and Zimbabwe demonstrate that access to water directly affects women’s time, livelihood opportunities, economic resource options, and health and wellbeing, and in turn, women’s time, livelihood, and economic resource options directly influence their economic engagement. While women did not discuss a direct influence of their health and wellbeing on their economic engagement, we expect that the reported negative impacts of water acquisition on health and well-being likely influence women’s economic engagement. Further, gender norms, life stage, drought, and seasonality exacerbate challenges to access water, hence constraining women’s economic opportunities.

### 4.1 Time

While health has remained the primary justification for improving water access(36), some have argued for improving water access specifically to reduce the time burden of water collection placed on women, potentially enabling them to engage in other opportunities, including but not limited to income generation(36, 37). Aligned with this argument, women across all four countries in our study discussed how the time burden of water collection constrained their ability to engage in economic opportunities. That said, research to date assessing water collection time and associations with economic engagement has been mixed, indicating both positive and negative associations of improved water access to women’s economic engagement. Ilahi and Grimard (2000) affirmed that improved water infrastructure increased the proportion of Pakistani women’s time that was allocated in income-generating activities and further reduced total work burden of female household members(38). Conversely, in Kyrgyzstan, improved water access amounted to an 80 minute increase in leisure time and a 90 minute increase in farm labor resulting in increased household cereal production, but not income-generating market labor(39). Likewise, in Benin, improved water supply resulted in a reduction in time for water collection by 41 minutes, resulting in women making additional trips, with no increase in market labor due to time gains(15). Furthermore, in their study on water access, women’s work, and child outcomes in nine countries across North and sub-Saharan Africa, South Asia, and the Middle East, Koolwal and Van De Walle (2013) reported no association between reduction in water collection time with women’s off-farm work and wage work in Yemen, Uganda, Nepal, Malawi, India, and Pakistan. In other countries, reduced water collection time led to less unpaid work and more leisure for women, but did not result in higher participation in market work or linked improved water access to economic engagement(40).

Consistent with other studies, our results also indicated that some women may not necessarily use the extra time on income generation; having time does not always translate to being able to participate in income-generating activities, since women may still lack skills, tools, or opportunities needed to find a job or start a business(39–41). Women may prioritize activities hierarchically based on their order of importance, which may leave economic engagement at the bottom of the hierarchy. In addition, as noted above, men in all countries dictated women’s activities, and women indicated that they required men’s permission to engage in economic activities, suggesting that women may have low agency to decide how to spend any time that is saved. Although improved access can alleviate water collection time needs, understanding women’s time needs, priorities, and time-use agency(42, 43) beyond water access is critical to ascertain the type, quality and sustainability prospects of activities women would engage in, including whether income generation is a viable option.

Beyond time needed to access water, women in Guatemala also discussed the time burden of engaging in committees. Women’s participation in water management is vital, since they are primary water collectors and users. Although water user committees with higher numbers of female members are more active, record high functionality of water sources, and ensure better access to water(44), women’s involvement in water committees, and the amount of time commitment, could deprive women of time to engage in other economic activities. A study in India found that an increase in women’s engagement in water management-related unpaid domestic services resulted in women’s lower labor force participation compared to men(45). Women in our study recognized that despite the time burden, their level of involvement in leadership roles within water committees would motivate them to fully participate, although other studies have indicated that major leadership roles in water committees are taken up by men, leaving women as participants and not actual contributors(46–48). As aligned with our findings, women may have no options but to contribute to community water programs if they are widowed, single or are the only available person to take up the role due to household complexities(49). Therefore, water programs must recognize intersectionality and the differences between women if the poorest and most disadvantaged women are to benefit(50), consider additional burden that water initiatives can have on women and men, and include strategies that can alleviate women from more unpaid work and benefit both men and women.

### 4.2 Livelihood opportunities

Consistent with other research, we found women’s engagement in agriculture and livestock production to be hindered by limited water access. Water access has been argued to be a ‘prerequisite’ for business and livelihood(36), and research shows that women’s livelihood-related water needs and access may be different and potentially more constrained compared to men in their own household(51, 52). Besides common water uses, including production such as agriculture and livestock production, and domestic purposes like bathing, washing, drinking, kitchen gardening, and watering animals, water needs for women and men differ based on crop preferences(53). These differences are also influenced by factors such as reduced availability of male family labor, for instance if women are widowed, divorced, or if husband migrated, and the transaction cost required to get water(54). These other factors may constrain women from engaging in livelihood opportunities even if there is water.

Furthermore, drought in Kenya and Zimbabwe, and seasonality in Guatemala and Honduras contributed to water scarcity, demonstrated through intermittent water supply defined as water supply that for a period of time is unavailable during rainy or dry season(55). Drought and seasonality continue to create significant challenges for access to water across different countries, phenomena that will possibly be worsened by climate change(56). Kenya and Zimbabwe have experienced reduced water tables and drying of available water sources as reported in other countries(57, 58). Beyond the importance of water for income generation, interviews with women in our study revealed how a depletion or lack of water can be detrimental to economic engagement. Specifically, women described how a reduction in water led to thinning and death of livestock and drying up of gardens and other crops. Taken together, our research and that of others suggests that programs aiming to improve water access in order to enable income generation through agriculture and livestock production must ensure the sustainability of water initiatives(59). Such programs could incorporate climate resilient WASH technologies as appropriate such as retention ponds or drip irrigation and sustainable catchment management practices that increase ground water recharge to address the increasing impact of climate change on water infrastructure(60). In addition, an in-depth understanding of the influence of intermittent water supply on agriculture and livestock productivity is needed.

### 4.3 Economic resource constraints

The payment for water at sources and costs incurred on installation and maintenance of water infrastructure were extensively mentioned by participants, and these factors could influence women’s economic engagement. Women described paying for water at the source, remitting monthly water bills, purchasing water storage vessels, and paying for water repairs and installation, which depleted their household income. Like other studies(23, 61), women in this study were engaged in agriculture, livestock rearing and other water intensive businesses, which increased their water demands and expenditure. Such expenses can reduce profit and viability of businesses. While improved water supplies can reduce direct costs of water, our findings, similar to another study, showed that the increased cost of operation and maintenance for water infrastructure (due to cost of pipes and labor) made it harder to access adequate water (62). Therefore, infrastructure improvements need to provide sufficient quantities of water and time savings, ideally without resulting in increased costs that can reduce women’s capital thereby reducing their ability to engage in business(15).

### 4.4 Health and wellbeing

Women in our study talked about how water collection impacted their health and well-being, and while they did not draw direct connections between their health and well-being and their economic engagement, previous research suggests a link likely exists and is worth further investigation. It is already widely acknowledged that women’s reproductive health is important for their economic empowerment, particularly women’s ability to control the timing and spacing of births during their years of peak economic productivity(63, 64). However, studies have more often treated health as an outcome of economic empowerment, rather than as a predictor of economic empowerment. For example, studies show that women’s economic rights to own and inherit property and have control over their earnings significantly improve their access to maternal health care, dietary diversity, and overall welfare(65–69). Women in our study highlighted how a lack of access to water influences their health and wellbeing including pain, fatigue, exhaustion, injury and others. Similarly, other research has found that water fetching is associated with poor musculoskeletal health like pain in the head, upper back, chest, hands, feet, and abdomen, and noted that these issues can be chronic (due to habitual head loading) or acute (due to environmental hazard-related injuries such as broken bones) (70, 71). Importantly, musculoskeletal conditions have been shown to profoundly affect economic wellbeing as well as physical and mental wellbeing and physical and emotional relationships(72). Evidence also suggests that water carriage impacts women’s health acutely. A cross-sectional study with 1001 women of reproductive age in Nepal found that women who did not receive help with water carriage had a 4.4 times higher odds ratio for uterine prolapse(73). Treatment of such injuries and conditions may be costly, potentially depleting household income and capital, and may render women physically unable to engage in economic activities. Musculoskeletal pains can often go undiagnosed(72), and may lead to long term physical disability, hence limiting opportunities and hindering women from fully participating in income-generating activities. An understanding of how health and wellbeing, influenced by water collection, impacts women’s economic engagement, is crucial for developing interventions that support women’s economic activities while addressing the challenges posed by water collection.

Like findings from other studies(11, 74, 75), existing gender norms in our study context reinforced the expectation that water collection be carried out by women. These expectations can be exacerbated by the deeply rooted patriarchal systems which support them and make it difficult to challenge these harmful norms(76). For instance, our study found that it was “shameful” for men to collect water, that women perceived men as “superior” and could not collect water, and that men were dissuaded from collecting water. A mixed-methods study that explored the gender differences in water collection in Makondo Parish in Uganda also found that traditional norms and stereotypes deemed it shameful, demeaning, and unusual for men to collect water (11). These gender norms reinforce the invisibility of the significant work of women, a finding also reported in a study which examined how the water fetching burden differed by gender among rural households in west region of Cameroon(77). Although culture and traditions can contribute to the reinforcement of harmful gender norms related to water collection(76), in our study, women discussed how men in Guatemala and Honduras could participate in water collection if they had time. This demonstrated that norms are malleable. Changing norms is necessary to relieve women of the burden of water collection, and to challenge the perception that it is expected and ‘free’ for women’s bodies to be used as a pathway or conduit to enable the flow of water to households(76).

### 4.5 Strengths and Limitations

This study has several strengths. Using qualitative data collection methods to understand barriers to economic engagement as they relate to water access was crucial in capturing diverse voices, experiences, and perspectives. Participants were able to explain how and why the identified barriers influenced women’s economic engagement, providing a comprehensive understanding of how these factors interacted within and across different settings. Men’s participation in FGDs and KIIs allowed for the exploration of how gender norms are operationalized and provided validation of women’s experiences, considering that male involvement can contribute to strengthening effectiveness of programming which address gender norms. Engaging local enumerators enabled building trust with participants as they knew the language and culture and shared lived experiences with the participants. Some limitations exist. Audio records were translated from local languages to English and during this process the meanings of some topics could have been lost or misconstrued. Furthermore, using community gatekeepers to recruit participants and selecting a convenience sample are limitations, as we may have missed key perspectives from diverse individuals who were not contacted. The season in which data was collected could have led participants to reference the most current season in their discussion of water access. Collecting data in other seasons could potentially illuminate other challenges that influence women’s economic empowerment.

## 5.0 Conclusion

Water is a prerequisite to economic engagement, especially in hard-to-reach low-resource settings. Access to water can enable livelihood opportunities, allow women to save or reallocate time, enable various economic resource options, and improve health and wellbeing, which can then facilitate women’s economic engagement. However, insufficient access to water can demand arduous water collection tasks, which can impact health by causing energy depletion, increasing risk of injury, and causing mental strain. These negative health impacts, coupled with time and opportunity costs, can constrain women’s abilities to engage in other facets of life, including economic engagement, which could provide well-being benefits to women and their families. While these results show how water fetching can influence women’s physical and mental health, further research is needed to understand how health and wellbeing can influence economic engagement, as many studies focus on health outcomes rather than the reverse. In addition, it is necessary to implement gender-transformative programming that aims to enhance women’s economic engagement by reducing the water-related barriers women face, while also addressing gender norms that prevent their full participation in economic activities.

## Supporting information

Supplemental checklist

Codebook; Tools

## Data Availability

All data produced in the present study are available upon reasonable request to the authors

## Acknowledgements

We are grateful to the participants of this study for their time and for sharing their experiences. We would like to thank the following partners and enumerators for their research collaboration. From Centro Universitario de Occidente, de la Universidad de San Carlos de Guatemala: Raul Bethancourt, Luis Alfredo López Cortez, Leonardo Cabrera López, Facundo Zacarías López Sánchez, Alex Fernando López Cortez, Zaida Anaí Vasquez García, Gladys Judith Vásquez García López, Vilma Rosario López Sánchez, Any Gabriela León Ramon, Heidy Maribel Baten Velásquez, Lucy Antonieta Ixmucané Capriel Herrera, Lesbia Beatriz Xiloj Hernandez, Ana Rosa León, José Joél Nolasco Yacabalquiej, Ingrid Johana Guachiac Gómez; from World Vision Guatemala: Enri Maldonado, Estuardo Gonzales; from Universidad Nacional Autónoma de Honduras: Rosaura Suyapa Rodriguez Funez, Ashly Michelle Méndez, Nastin Nahomy Tilguant Ávila, Brenda Paola Cruz, Yesi Karolina Murillo Vega, Jorge Luis Cálix Barahona, Dulce Milagro Sevilla Sosa, Wilber Adonay Ávila Valladares, Néstor Fabricio Dávila González, Pedro Alejandro Hernández; from St. Paul’s University, Kenya: Peter Koome, Petronilla Andiba Otuya, Lesoito Jeremiah Christopher, Leshaule Benedicto Alois, Patricia Lekiliyo, Lechalote Ben Ljania, Nasieku Lolchuragi, Lekalantula Raina Deborah, Glory Karimi Kiriinya, Lekonte Salome Mary, Lelikoo Naanyu Sarah, John Kalasinga Lerosion; from World Vision Kenya: Kevin Muche; and from Datalyst Africa: Molly Manyonganise, Eve Nyemba, Sunga Mzeche, Nyasha Kamwire, Luckness Zimanyiwa, Sibonisiwe Mpofu, Tapiwa Mapfumo, Kudzai Zibgwi, Pricilla Moyo, Gladys Mhaka, Sithokozile Sibanda, Gugulethu Mnkandhla. Lastly, we would like to thank Jorge Beteta for his translation services.

