## Supplemental checklist for "“If we have water, we have money”: A qualitative investigation of the role of water in women’s economic engagement in Guatemala, Honduras, Kenya, and Zimbabwe"

Inclusivity in global research

PLOS’ policy on inclusivity in global research aims to improve transparency in the reporting of research performed outside of researchers’ own country or community and ensures that PLOS publications reporting global research adhere to high standards for research ethics and authorship. Authors of relevant research articles may be asked to complete the questionnaire below, which outlines ethical, cultural, and scientific considerations specific to inclusivity in global research. This questionnaire may be requested when researchers have travelled to a different country to conduct research, if research uses samples collected in another country, research with Indigenous populations or their lands, or if research is on cultural artefacts. Researchers travelling to another country solely to use laboratory equipment will not normally be required to complete the questionnaire. However, the questionnaire can be requested at the journal’s discretion for any submission – if you have been requested to complete this questionnaire by the PLOS journal you submitted to, please do so.

Please complete the questionnaire below and include this as a Supporting Information file with your manuscript. Note that if your paper is accepted for publication, this checklist will be published with your article in the supporting information files. Please ensure that you reference the checklist in the main body of your manuscript. We suggest adding a subsection ‘Inclusivity in global research’ to your Methods section and adding the following sentence: “Additional information regarding the ethical, cultural, and scientific considerations specific to inclusivity in global research is included in the Supporting Information (SX Checklist)”

The questions have been designed to be applicable to a wide range of study types, and there are subsections for both human subjects research and non-human subjects research. If any of the questions are not relevant to your research please mark them as “N/A” as appropriate.

**Ethical considerations, permits and authorship**

*This section is applicable to all research types.*Provide details as to who granted permissions and/or consent for the study to take place in the Methods section of your manuscript. This should include the names of **all**

ethics boards, governmental organizations, community leaders or other bodies that provided approval for the study. If individuals provided approval refer to these people by their role or title but do not list their name(s).

Reported on page number: **Reported on page 11 of the manuscript: This study was considered exempt by the Institutional Review Board committee of Emory University, Atlanta, Georgia, USA (IRB00005955), and was approved by the Comité de Ética del Centro Universitario de Occidente, de la Universidad de San Carlos de Guatemala in Guatemala (Acta 1.23 C.E. DICUNOC); Universidad Nacional Autónoma de Honduras (CEIFCS-2023-P18) in Honduras; St. Paul’s University - Institutional Scientific Ethics Committee (ERB No. 38), the National Commission for Science, Technology and Innovation (NACOSTI/P/23/27117) in Kenya; and the Medical Research Council of Zimbabwe (MRCZ/A/3054) in Zimbabwe.**

If there were any deviations from the study protocol after approval was obtained please provide details of these changes in the Methods section of your manuscript.

Reported on page number: **There were no major deviations from study protocol for this study**

Did this study involve local collaborators that are residents of the country where the research was conducted or members of the community studied? If you do not have any authors from said communities, please provide an explanation for this below.

**This study involved local collaborators in Guatemala, Honduras, Kenya, and Zimbabwe. Our co-authors include collaborators from each of our country-based data collection and project implementation partners.**

Everyone listed as an author should meet PLOS’ criteria for authorship and all individuals who meet these criteria should be included in the author byline, rather than the acknowledgements. For further information please see the journal’s Authorship Policy.

**Human subjects research (e.g. health research, medical research, cross-cultural psychology)**

Did you obtain written informed consent from a representative of the local community or region before the research took place? How did you establish who speaks for the community? Details of written informed consent obtained from study participants should be reported separately in the Methods section of your manuscript.

**Before research took place, written informed consent was obtained from district government in Zimbabwe per local Zimbabwe IRB requirements; verbal consent was obtained from community leaders in Guatemala, Honduras, Kenya, and Zimbabwe. In addition to the local data collection teams, World Vision had an initial role in establishing who spoke for community consent as they work with the community leadership for project implementation**

How did members of the local community provide input on the aims of the research investigation, its methodology, and its anticipated outcome(s)?

**Members of the local communities did not provide input on the aims of the research investigation, its methodology, and its anticipated outcomes. However, World Vision country partners, who work closely with members of the local communities, provided input on the aims of the research investigation. Similarly, the country-based data collection teams provided input on methodology and assisted revising and translating the qualitative tools for local context and participant understanding.**

When engaging with the local community, how did you ensure that the informed consent documents and other materials could be understood by local stakeholders?

**Informed consent documents were written in local languages (Guatemala: Kiche, Mam, and Spanish; Honduras: Spanish; Kenya: Samburu; and Zimbabwe: Shona and Ndebele) and enumerators read the consent documents in the local language preferred by each participant. All participants provided informed consent. In Guatemala, Honduras, and Kenya, participants consented verbally, and enumerators signed the consent form to affirm consent. In Zimbabwe, participants provided signed consents. Following informed consent, enumerators conducted data collection in the local language preferred by each participant****.**

Will the findings of the research be made available in an understandable format to stakeholders in the community where the study was conducted (e.g. via a presentation, summary report, copies of publications, etc.)? Please provide details of how this will be achieved.

**The findings of the research are anticipated to be shared with communities where the study was conducted in Guatemala, Honduras, Kenya, and Zimbabwe. World Vision country offices are planning to hold meetings that invite community members and leaders, government stakeholders, and program leaders in each of the four countries. Presentation formats will vary depending on the location, but all will aim to be conducted in an understandable manner (e.g., using graphics to describe findings) and in local dialects (e.g., Samburu in Kenya).**

**Non-human subjects research using specimens/ animals collected as part of the study, or those housed in archival collections. Examples include archaeology, paleontology, botany and zoology.**

Did the permission you obtained from a local authority to perform the study include an agreement on access to outputs and benefit sharing? This may include procedures to enable fair distribution of the benefits and resources arising from the research performed. Please include any details of Prior Informed Consent and Benefit Sharing Agreements obtained. These may be required by field-specific regulations, for example the Convention on Biological Diversity (CBD) and the associated Nagoya Protocol.

**N/A**

If the material used in your study was imported, please A) provide the year it was imported and B) indicate whether permits were obtained to import/export the materials used, C) provide details of any permits obtained. If this information is not available, please indicate this.

**N/A**

If you used archival specimens, please state how the material used in your study was acquired by the institute it is held in and provide details of any permits obtained for the original excavations/ sample collection. If this information is not available, please indicate this.

**N/A**

How was the potential cultural significance of the materials collected in your study to local communities considered in your research design? Were Indigenous peoples and/or local researchers and institutions involved with archaeological excavations / collection of specimens? If so, please provide a description of their involvement.

**N/A**

If your manuscript includes photographs of human remains please indicate whether authors obtained permission from descendants or affiliated cultural communities to do so.

**N/A**
