## Supplementary material for "“If we have water, we have money”: A qualitative investigation of the role of water in women’s economic engagement in Guatemala, Honduras, Kenya, and Zimbabwe": Codebook; Tools: S1 Tools.docx

**S1 Tools.** Data collection tools for participant demographic information, focus group discussion with women, focus group discussion with men, and key informant interviews

**S1a Tools.** Participant Demographic Information

| **A. Activity information** | | | |
| --- | --- | --- | --- |
| **A001** | Participant ID#: __________________________________ | **A005** | Date: (y/d/m) __ __ __ __ / *__ _**_ / __ __* |
| **A010** | Community Name: __________________________________ | **A015** | Community ID#: ____________________________________ |
| **A017** | **Select activity person will participate in:** | ☐ 1. Key Informant Interview  ☐ 2. Water Journey Go-Along Interview  ☐ 3. FGD with Women  ☐ 4. FGD with Men | |
| **A18** | **If there is more than one activity of this type today, indicate activity number:** | ☐ 1. Not applicable; only activity of this kind here today  ☐ 2. Activity number: **__ __** | |
| **A020** | **Activity Start time:** __ __ : __ __ pm / am | **A025** | **Activity End time:** __ __ : __ __ pm / am |
| **A030** | **Person Filling** **form:__________________** | **A040** | **Consent Obtained:** ☐ 1. Yes ☐ 2. No |

| **B. Participant Demographic Information** | | | |
| --- | --- | --- | --- |
| **D01** | **Participant Gender**  ☐ 1. Woman ☐ 2. Man | **D02** | **Participant Age __ __**  *(If younger than 18, participant ineligible. End.)* |
| **D03a** | **What is your marital status?**  ☐ 1. Single, never been married  ☐ 2. Unmarried, but have partner  ☐ 3. Married  ☐ 4. Separated  ☐ 5. Divorced  ☐ 6. Widowed | **D03b** | **What type of family structure do you have? [Kenya]**  ☐ 1. Monogamy  ☐ 2. Polygamy |
| **D04** | **What is the highest level of school you completed?**  ☐ 1. Never attended school  ☐ 2. Completed some Primary  ☐ 3. Completed Primary  ☐ 4. Completed Secondary  ☐ 5. Completed schooling above Secondary | | |
| **D05** | **How many people (including yourself) live in your household? __ __**  [Usually number of people sharing meals] | **D06** | **How many children live in your household?**  **__ __**  [This is the total number under age 18.] |
| **D07a** | **Some people take up jobs for which**  **they are paid in cash or kind. Others sell things, have a small business or work on the family farm or in the family business. In the last** **30 days, have you done any of these things or any other work?** ☐ 1. Yes ☐ 2. No | **D07b** | ***If participant has engaged in work in the past 30 days (D07a=yes):***  **What are the activities you have done for work?** (write answer) |
| **D08a** | ***If married,* In the last** **30 days, has your spouse [husband/wife] done any of these things or any other work?**  ☐ 1. Yes ☐ 2. No | **D08b** | ***If participant’s spouse has engaged in work in the past 30 days (D08a=yes):***  **What are the activities your spouse has done for work?** (write answer) |
| **D09a**  **D09b** | **What is the primary source of DRINKING water used by members of the household in the past 7 days / week?**  **What is the primary source of water FOR OTHER NEEDS used by members of the household in the past 7 days / week?**  **(includes for cooking, bathing, washing clothing, etc.)** | **D09a D09b**  **Drinking Other Uses**  ☐ ☐ 01. No other source used  ☐ ☐ 10. Piped Water  ☐ ☐ 21. Tube well/Borehole  ☐ ☐ 31. Protected Dug Well  ☐ ☐ 32. Unprotected Dug Well  ☐ ☐ 41. Protected Spring  ☐ ☐ 42. Unprotected Spring  ☐ ☐ 51. Rainwater  ☐ ☐ 61. Tanker Truck  ☐ ☐ 71. Cart with small tank  ☐ ☐ 72. Water Kiosk  ☐ ☐ 81. Surface Water *  ☐ ☐ 91. Packaged bottled water  ☐ ☐ 92. Packaged Sachet water  ☐ ☐ 96. Other_____________ *River, dam, lake, pond, stream, canal, irrigation channel | |
| **D10a** | **Where is the household’s main drinking water source?**  ☐ 1. Source in dwelling  ☐ 2. Source in yard/plot  ☐ 3. Source elsewhere (beyond yard/plot) | **D10b** | **How long does it take for members of your household to go to the drinking water source, get water, and come back?**  ☐ 1. __ __ __ minutes  ☐ 88. Do not know  ☐ 99. Not applicable; do not collect drinking water /source on property |
| **D10c** | **How many days in a week does your household collect water from the primary drinking water source?**  __ __  Enter 1-7 for number of days per week  Enter 99 if not applicable  Enter 88 if do not know | **D10d** | **Who is *primarily responsible* for collecting drinking water for the household?**  ☐ 1. Respondent  ☐ 2. Adult woman (age 18 or over)  ☐ 3. Girl (under age 18)  ☐ 4. Adult man (age 18 or over)  ☐ 5. Boy (under age 18)  ☐ 99. Not applicable; do not collect drinking water |
| **D11** | **In the last month, has there been any time when your household did not have** **sufficient quantities of drinking water when needed?**  ☐ 1. Yes, at least once  ☐ 2. No, Always sufficient  ☐ 88. Do not know | **D11a** | ***If the source for other uses is different than the drinking water source,***  **Where is the household’s main water source for *other uses*?**  ☐ 1. Source in dwelling  ☐ 2. Source in yard/plot  ☐ 3. Source elsewhere (beyond yard/plot) |
| **D11b** | ***If the source for other uses is different than the drinking water source,***  **How long does it take for members of your household to go to the water source for other uses, get water, and come back?**  ☐ 1. __ __ __ minutes  ☐ 88. Do not know  ☐ 99. Not applicable; do not collect drinking water /source on property | **D12** | **Who is *primarily responsible* for collecting water for other uses for the household?**  ☐ 1. Respondent  ☐ 2. Adult woman (age 18 or over)  ☐ 3. Girl (under age 18)  ☐ 4. Adult man (age 18 or over)  ☐ 5. Boy (under age 18)  ☐ 99. Not applicable; do not collect water for other uses |
| **D13a** | **Does your household have access to a toilet facility?**  ☐ 1. Yes, in dwelling  ☐ 2. Yes, in yard/plot  ☐ 3. Yes, elsewhere (beyond yard/plot)  ☐ 4. No household access to a toilet facility | **D13b** | **If your household has access to a toilet facility, do you share it**?  ☐ 1. No, not shared with other households  ☐ 2. Yes, shared with specific households  ☐ 3. Yes, shared with public/community  ☐ 99. Not applicable; no access to a toilet facility |

**S1b Tools.** Focus Group Discussion Guide for Women

**Guide: Focus Group Discussion with Women**

**Primary Goal:** To understand women’s perceived barriers and facilitators to economic empowerment activities.

**Specific Objectives**

- To understand women’s current involvement in economic activities in the community (e.g., income generation, saving, taking loans, making purchases), including examples of how they are involved.
- To understand perceptions of women’s involvement in economic activities, including what participants think women perceive and what men perceive.
- To understand perceived barriers and facilitators to women’s involvement in economic activities, with specific attention to program elements (e.g., gender norms, access to savings/loans, skills to manage money, water access, etc.).
- To understand participant perceptions of if and how improvements to water access and availability may influence women’s ability to participate in economic activities.

| **Guide: Focus Group Discussion with Women** | | | |
| --- | --- | --- | --- |
| **A010.** | Community Name: _____________________ | **A015.** | Community ID#: ________________________ |
| **A020.** | Activity Start time: __ __ : __ __ pm / am | **A025.** | Activity End time: __ __ : __ __ pm / am |
| **A055.** | Date: (y/d/m) __ __ __ __ / *__ __ / __ __* | **A040.** | Consent Provided by All: ☐ 1. Yes ☐ 2. No |
| **A045.** | Recorder ID:_________________________ | **A046.** | Recording #_______________________ |
| **A030.** | Facilitator: __________________________ | **A035.** | Note Taker: __________________________ |
| **Introduction** | | | |
| **We are gathering today because we are interested in learning more about experiences women like you have. We are specifically interested in learning if women in this community do any activities to earn, save, loan and spend money, what those activities are. We are also interested to understand what may make it hard for women to do these activities, and what could make it easier. To make you more comfortable, you do not need to share what you experience specifically. You can discuss what you know may be concerns for other women in the community like you.**  **GROUND RULES / GUIDELINES**   1. Please respect what others say. Everyone deserves a chance to speak. Please do not interrupt when someone else is talking. If people speak over each other, it will be hard to understand. 2. If you begin talking when another is speaking, I will ask you to wait or I may hold up my hand to tell you to stop. But, I will get back to you when the person is finished. 3. Please do not have conversations with those next to you as it will interfere with the discussion. 4. This is a discussion, so please listen to what others are saying so that you can add your thoughts. You are welcome to share agreement and disagreement. There are no right or wrong answers. | | | |
| **Warm-up**  *Note for facilitator (do not read out loud): This question is designed to get everyone talking right at the beginning. If you feel people are still shy, we can have them go around and share something more.* | | | |
| **1. WARM UP**  **First, we would like to start by having everyone share something about themselves. You do not need to say your name. Please go around and share. You can tell us about your family, your favorite food, or anything you like.** | | | |
| **Key Questions** | | | |
| **Thank you all for sharing. We will now start talking about your experiences.** | | | |

| A. **Economic activities** | |
| --- | --- |
| **1. What are some of the economic activities that women are engaged with in this community?**  *-Allow participants to provide answers before asking about specific activities. For each activity noted, ask them to provide details. ‘Can you tell me more about that?’*  *-To encourage them to provide more examples, you can ask ‘anything else?’ ‘What else?’ etc.* | *-Probe about the following if not already noted in participant responses:*  - women’s income generation (for herself; for others in the household (e.g. post-harvest processing of crops that are then sold by her husband);  - activities that may not generate income but produce goods that can be bartered or exchanged;  -activities that are paid in-kind rather than in cash;  -savings groups  -loan groups  -economic or business education/learning |
| **2. What are some of the factors that have enabled or helped women to participate in economic activities in this community?** | Probe:  Available time?  Opportunities?  Cultural norms?  Social support? (e.g. from household members, community, etc.,);  Access to resources such as training and credit?  Access to resources such as raw materials (water, clay, seeds)  Economic factors- markets, loans, banks  Geography/environment?  Government policies/ licensing? |
| **3. What are some of the barriers that have made it hard for women to participate in economic activities in this community?** | Probe:  Available time?  Competing responsibilities?  Opportunities?  Cultural norms?  Social support? (e.g. from household members, community, etc.,);  Access to resources such as training and credit?  Economic factors- lack of markets, banks, loans, inflation?  Geography/ environment?  Government policies/ licensing?  Specific skills required to be able to participate in economic activities? |
| **4. What do women in this community think of other women who engage in the economic activities you noted?** | Probes:  Why do they feel this way?  Support/Think positively?  Do not support/ Think negatively? |
| **5. What do MEN in this community think of other women who engage in the economic activities you noted?** | Probes:  Why do they feel this way?  Support/Think positively?  Do not support/ Think negatively? |
| **6. If women in this community generate income, who decides how the money is used?** | Probes:  -What do women most often use money for if they can use it?  -How different from what men use money for?  - Decision making process on how and what they are going to use the money (Do women participate meaningfully in making decision?) |
| **7. What would women in this community think if there was a program:**  a. For women to learn to save money?  b. For women to learn how to make money/start a small business?  c. For women to take a loan?  d. For women to learn about child health? | Probe:  What would men think of each of these activities for women? |
| **We will now discuss water access and collection in your community.** | |
| **B. Water** | |
| 1**. Please describe how and where people typically get water for their household in this community?** | Probe:  -How long does it take?  -Who is usually responsible?  -Where?  -Source?  -Near/Far from household?  -Different location or frequency for different needs?, seasons? |
| **2. What challenges do women face collecting water for their household or doing water-related work (e.g. laundry, cleaning)?** | Probe:  -Risk?/ Safety?  -Terrain?  -Injury?  -Labor? |
| **3. What benefits do women experience when collecting water for their household or doing water-related work?** | Probe:  -Social time with other women  -Spending time outside the house |
| **4. How do you think the lives of women would change if you had better access to water?** | Probe:  - time to engage in other activities  - leisure/ rest,  -energy/health  - income generating activities  - time with family? |

**S1c Tools.** Focus Group Discussion Guide for Men

Guide: Focus Group Discussion with Men

**Primary Goal:** To understand women’s perceived barriers and facilitators to economic empowerment activities.

**Specific Objectives**

- To understand women’s current involvement in economic activities in the community (e.g., income generation, saving, taking loans, making purchases), including examples of how they are involved.
- To understand perceptions of women’s involvement in economic activities, including what participants think women perceive and what men perceive.
- To understand perceived barriers and facilitators to women’s involvement in economic activities, with specific attention to program elements (e.g., gender norms, access to savings/loans, skills to manage money, water access, etc.).
- To understand participant perceptions of if and how improvements to water access and availability may influence women’s ability to participate in economic activities.

**FGD with Men**

| **Guide: Focus Group Discussion with Men** | | | |
| --- | --- | --- | --- |
| **A010.** | Community Name: _____________________ | **A015.** | Community ID#: ________________________ |
| **A020.** | Activity Start time: __ __ : __ __ pm / am | **A025.** | Activity End time: __ __ : __ __ pm / am |
| **A055.** | Date: (y/d/m) __ __ __ __ / *__ __ / __ __* | **A040.** | Consent Provided by All: ☐ 1. Yes ☐ 2. No |
| **A045.** | Recorder ID:_________________________ | **A046.** | Recording #_______________________ |
| **A030.** | Facilitator: __________________________ | **A035.** | Note Taker: __________________________ |
| **Introduction** | | | |
| **We are gathering today because we are interested in learning more about your thoughts on women’s roles and work in the community. We are specifically interested in learning if women in this community do any activities to earn, save, loan and spend money, what those activities are. We are also interested to understand what may make it hard for women to do these activities, and what could make it easier. We are also interested in learning about water access in the community. To make you more comfortable, you do not need to share what women in your household experience. We are interested in perceptions for the entire community.**  **GROUND RULES / GUIDELINES**   1. Please respect what others say. Everyone deserves a chance to speak. Please do not interrupt when someone else is talking. If people speak over each other, it will be hard to understand. 2. If you begin talking when another is speaking, I will ask you to wait or I may hold up my hand to tell you to stop. But, I will get back to you when the person is finished. 3. Please do not have conversations with those next to you as it will interfere with the discussion. 4. This is a discussion, so please listen to what others are saying so that you can add your thoughts. You are welcome to share agreement and disagreement. There are no right or wrong answers. | | | |
| **Warm-up**  *Note for facilitator (do not read out loud): This question is designed to get everyone talking right at the beginning. If you feel people are still shy, we can have them go around and share something more.* | | | |
| **1. WARM UP**  **First, we would like to start by having everyone share something about themselves. You do not need to say your name. Please go around and share. You can tell us about your family, your favorite food, or anything you like.** | | | |
| **Key Questions** | | | |
| **Thank you all for sharing. We will now start talking about your experiences.** | | | |

| **Main questions** | **Sub questions and probes** |
| --- | --- |
| **Water access and use** | |

| **Main questions** | **Sub questions and probes** |
| --- | --- |
| **1. What are some of the economic activities that women are engaged with in this community?**    *-Allow participants to provide answers before asking about specific activities. For each activity noted, ask them to provide details. ‘Can you tell me more about that?’*    *-To encourage them to provide more examples, you can ask ‘anything else?’ ‘What else?’ etc.* | *-Probe about the following if not already noted in participant responses:*    - women’s income generation (for herself; for others in the household (e.g. post-harvest processing of crops that are then sold by her husband);    - activities that may not generate income but produce goods that can be bartered or exchanged;    -activities that are paid in-kind rather than in cash;    -savings groups    -loan groups     - -economic or business education/learning |
| **2. What are some of the factors that have enabled or helped women to participate in economic activities in this community?** | Probe:  Available time?  Opportunities?  Cultural norms?  Social support? (e.g. from household members, community, etc.,);  Access to resources such as training and credit?  Access to resources such as raw materials (water, clay, seeds)  Economic factors- markets, loans, banks  Geography/ environment?   - Government policies/ licensing? |
| **3. What are some of the barriers that have made it hard for women to participate in economic activities in this community?** | Probe:  Available time?  Competing responsibilities?  Opportunities?  Cultural norms?  Social support? (e.g. from household members, community, etc.,);  Access to resources such as training and credit?  Economic factors- lack of markets, banks, loans, inflation?  Geography/ environment?  Government policies/ licensing?   - Specific Skilled required to be able to participate in economic activities? |
| **4. What do men in this community think of other women who engage in the economic activities you noted?** | Probes:  Why do they feel this way?  Support/Think positively?   - Do not support/ Think negatively? |

| **5. If women in this community generate income, who decides how the money is used?** | Probes:  - Decision making process on how and what they are going to use the money (Do women participate meaningfully and make decisions?)  -What do women most often use money for if they can use it?  -How different from what men use money for? |
| --- | --- |
| **7. What would men in this community think if there was a program:**    a. For women to learn to save money?  b. For women to learn how to make money/start a small business?  c. For women to take a loan?  d. For women to learn about child health? | Probe:    Do you think women want to participate in these activities? How would it affect f |
| **We will now discuss water access and collection in your community.** | |
| **B. Water** | |
| **1. Please describe how and where people typically get water for their household in this community?** | Probe:  -How long does it take?  -Who is usually responsible?  -Where?    -Source?    -Near/Far from household?    -Different location or frequency for different needs?, seasons? |
| **2. What challenges do people face collecting water for their households?** | Probe:  -Risk?/ Safety?  -Terrain?  -Injury?  -Labor? |
| **3. How do you think the lives of people in this community would change if you had better access to water?** | Probe:  For women? For men?  - time to engage in other activities  - leisure/ rest,  -energy/health  - income generating activities  - time with family? |

**S1d Tools.** Key Informant Interview Guide

**Key Informant Interview guide**

| **Guide: Key Informant Interview Guide** | | | |
| --- | --- | --- | --- |
| **A010.** | Community Name: _____________________ | **A015.** | Community ID#: ________________________ |
| **A020.** | Activity Start time: __ __ : __ __ pm / am | **A025.** | Activity End time: __ __ : __ __ pm / am |
| **A055.** | Date: (y/d/m) __ __ __ __ / *__ __ / __ __* | **A040.** | Consent Provided: ☐ 1. Yes ☐ 2. No |
| **A045.** | Recorder ID:_________________________ | **A046.** | Recording #_______________________ |
| **A030.** | Facilitator: __________________________ | **A035.** | Note Taker: __________________________ |

**Objectives**

- To understand the community leader’s experiences and perspectives related to water access and use in the community
- To understand the perceptions of community leaders on women’s economic engagement

| **Main questions** | **Follow up questions and probes** |
| --- | --- |
| **Introduction** | |
| 1. What is your role in this community? | a. Probe: Roles/responsibilities related to water |
| **Water access and use** | |
| 2. What are the community members experiences and perceptions of collecting water? | a. Among households in this community, which household members usually have responsibility for water collection?    i. Why? b. Where do people collect water from? c. Are there specific areas designated for specific people at the source?    i. Why? |
| 3. How are water sources managed in this community? | a. Who is responsible?    i. Community water committee    ii. Private/individual    iii. Women b. How are those responsible selected? c. Who decides on who to be included in these committees?    i. Why? |
| 4. How are women involved in management of water sources in this community? | a. Member of water management committee b. Leadership role in water management committees c. Contribution in decision making on water related issues |
| 5. What are some challenges that this community has encountered related to water? | a. Specific sub-groups within this community?    i. Households that have less access to a water source due to physical geography / infrastructure or social reasons    ii. Women    iii. School children b. If not mentioned, probe: Distance to the source i. Safety of water ii. Security e.g., banditry, wild animals iii. Prioritization of water (human vs animals) iv. Seasonality v. Cost vi. Operations and maintenance of water infrastructure (e.g. pumps) |
| 6. How have people addressed the challenges with water in this community? | Probe: Individual vs collective strategies |
| **Economic empowerment activities** | |
| 1. What economic activities are common in this community? | Probe:   a. For women b. For men |
| 2. Who decides which economic activity households participate in? Why? | Probe:   a. Self, men, women b. What is the perception of the community on women engaging in economic activities? |
| 3. What types of programs would you like to see in your community that would enable women to engage in economic activities? | Probe (don’t suggest these options: let the participant speak):   a. Savings groups b. Water committees that include women c. Nurture groups |
| 4. What are the facilitators to women’s involvement in economic activities in this community? | Probe:   a. Training b. Financial access c. Geography d. Access to markets e. |
| 5. What are some of the challenges to involvement in economic activities in this community? | Probe:   a. Sociocultural norms b. Geography c. Poverty d. Lack of markets   How have these challenges been addressed? |
| **Decision making related to water and economic activities** | |
| 1. Who makes decisions about community water resources in this community? | Probe on:   a. Who is involved, women, men, selected members of the community? Why? b. Where to start/ initiate community water source? c. What determines whether an individual qualifies for holding leadership positions in community owned projects? d. What is women’s contribution in the management of water sources?    i. Are women involved in leadership roles in water source management?    ii. Which positions?    iii. Who decides when women are involved |
| 2. Who makes decision on whether women engage in economic activities in this community? | a. Which economic activities are women commonly engaged in? b. Who decides how to spend resources in the household? Why? |
| 3. What would you recommend to ensure that women are more engaged in economic empowerment roles in this community? | a. Cultural shift b. Mindset change trainings/sessions |
